# Multinational evaluation of anthropometric age (AnthropoAge) as a measure of biological age in the USA, England, Mexico, Costa Rica, and China: a population-based longitudinal study

**DOI:** 10.1101/2024.07.09.24310149

**Authors:** Carlos A. Fermín-Martínez, Daniel Ramírez-García, Neftali Eduardo Antonio-Villa, Jerónimo Perezalonso Espinosa, Diego Aguilar-Ramírez, Carmen García-Peña, Luis Miguel Gutiérrez-Robledo, Jacqueline A. Seiglie, Omar Yaxmehen Bello-Chavolla

## Abstract

**OBJECTIVE:** To validate AnthropoAge, a new metric of biological age (BA), for prediction of all-cause mortality and age-related outcomes and characterize population-specific aging patterns using multinational longitudinal cohorts.

**METHODS:** We analyzed harmonized multinational data from the Gateway to Global Aging, including studies from the US, England, Mexico, Costa Rica, and China. We used body mass index and waist-to-height ratio to estimate AnthropoAge and AnthropoAgeAccel in participants aged 50-90 years old as proxies of BA and age acceleration, respectively. We compared the predictive capacity for all-cause mortality of AnthropoAge and chronological age (CA) using Cox models, described aging trends in all countries and explored the utility of longitudinal assessments of AnthropoAgeAccel to predict new-onset functional decline and age-related diseases using generalized estimating equations (GEE).

**FINDINGS:** Using data from 55,628 participants, we found AnthropoAge (c-statistic 0.772) outperformed CA (0.76) for prediction of mortality independently of comorbidities, sex, race/ethnicity, education, and lifestyle; this result was replicated in most countries individually except for Mexico. Individuals with accelerated aging had a ∼39% higher risk of death, and AnthropoAge also identified trends of faster biological aging per year. In longitudinal analyses, higher AnthropoAgeAccel values were independently predictive of self-reported health deterioration and new-onset deficits in basic/instrumental activities of daily living (ADL/IADL), diabetes, hypertension, cancer, chronic lung disease, myocardial infarction, and stroke.

**CONCLUSIONS:** AnthropoAge is a robust and reproducible BA metric associated with age-related outcomes. Its implementation could facilitate modeling trends of biological aging acceleration in different populations, although recalibration may enhance its utility in underrepresented populations such as individuals from Latin America.

## INTRODUCTION

Aging is a highly heterogeneous and complex process that leads to a progressive deterioration in multiple body systems, ultimately leading to disease, disability and death^1,2^. Although chronological age (CA) is a key risk factor for these outcomes, it is often unable to capture the inherent variability of the aging process, and numerous efforts have been made to quantify this heterogeneity and predict age-related outcomes. Biological age (BA) metrics, often called aging clocks^3^, are measures expressed in units of years that combine numerous aging biomarkers and have been proposed as an alternative to CA as they capture changes in biological systems more precisely, better reflecting the true status of health and biological decline of an individual^4,5^. First generation aging clocks are trained to predict CA from individual biomarkers, while second generation aging clocks are trained to predict age-related outcomes, such as mortality risk and comorbidities^6^; finally, third generation aging clocks are able to model the pace of aging by capturing longitudinal changes in aging biomarkers^7,8^. Recent evidence suggests that BA clocks capture unique aging processes, which reflect trajectories of functional, cognitive, and physical decline in an individual and may be superior to CA alone^9–11^. Despite this evidence, the utility of BA metrics remains controversial, as many other frameworks closely related to biological aging have been widely implemented in clinical geriatric assessments for years^12^. Nonetheless, the development of aging clocks continues to be relevant as currently, there is a lack of reproducible, precise, and simple methods that can serve as surrogates for the aging rate of an individual, which would allow to study population aging patterns, evaluate interventions aimed at healthy aging, and even uncover new biological mechanisms that explain the heterogeneity of this process^13,14^. Most BA measures have been developed using blood methylation data, while more recent approaches have incorporated a wide array of omics to integrate data of different system levels into the assessment of aging^15^.

However, due to the complexity of implementing omics-based measures in epidemiological studies, the application of BA to measure aging at a population-level remains limited^16^.

Our team recently developed AnthropoAge, which is a non-invasive alternative to measure BA using anthropometry to capture body composition aging and mortality risk^17^, thus representing an accessible and potentially useful aging biomarker to assess population aging^5,16^. Although AnthropoAge was developed using nationally representative data of participants with diverse ethnical and racial backgrounds, this may not fully capture the wide range of diversity in body composition across ethnicities since all participants from the original study were from the U.S. Whether our metric yields an adequate performance in diverse populations across different geographic contexts has not yet been explored. Furthermore, whether repeated assessments of AnthropoAge may be useful to detect functional and health outcomes has not been examined either. Leveraging longitudinal data from adults ≥50 years from the United States, England, Mexico, Costa Rica, and China, the main objective of this study is to validate AnthropoAge as a BA metric predictive of all-cause mortality, health deterioration and functional deficits in diverse populations, and to use it to characterize trends of population aging.

## METHODS

### Data access

For this population-based, multinational, longitudinal study, we used harmonized datasets from the Gateway to Global Aging Data platform (G2A, https://g2aging.org/), which gathers studies from multiple countries designed to be comparable with each other; these studies are longitudinal, multidisciplinary and nationally representative of the older population from its respective countries. We included individuals with complete data on core interviews, anthropometry, and linked mortality from the following cohorts: US Health and Retirement Study (HRS) , the English Longitudinal Study of Ageing (ELSA)^19^, the Mexican Health and Aging Study (MHAS)^20^, the Costa Rican Longevity and Healthy Aging Study (CRELES)^21^, and the China Health and Retirement Longitudinal Study (CHARLS)^22^. An overview of datasets, timepoints, and references to additional documentation for each study is provided in **Supplementary Methods.**

### Subsets of anthropometric data

In addition to demographic, socioeconomic and health-related questionnaires, a subset of participants in each study underwent further biochemical and physical measurements (including anthropometry) that were conducted at selected survey cycles; thus, the following subsets were the focus of our study:

- For HRS, participants were eligible for anthropometric measurements every two waves from waves 8 to 14 (2006 to 2018) among each half-sample of respondents (i.e., one half of participants has anthropometric data for waves 8, 10, 12 and 14, while the other half has this data for waves 9, 11 and 13)^18^.
- For ELSA, all participants from waves 2, 4, and 6 (2004, 2008, and 2012) were eligible for anthropometric measurements^19^.
- For MHAS, a random subsample of participants from waves 1 and 2 (2001 and 2003) was eligible for anthropometric measurements; however, measurements were restricted to the full cohort of only four states (one highly urbanized state, one with high-migration, one relatively poor and one with a high prevalence of diabetes) in participants from wave 3 (2012)^20^.
- For CRELES, participants from all waves were eligible for anthropometric measurements; however, we focused exclusively on participants from waves 1-3 (2005, 2007, 2010), which comprised the original pre-1945 cohort^21^ (see details in **Supplementary methods**).
- For CHARLS, participants from waves 1, 2 and 3 (2011, 2013 and 2015) were eligible for anthropometric measurements^22^.

With this data, we used measured height, weight, and waist circumference (self-reported anthropometry was not used) to calculate the body-mass index (BMI, obtained by dividing weight in kilograms by the square of height in meters) and the waist-to-height ratio (WHtR, dividing waist in centimeters by height in centimeters) of each participant. An overview of measuring techniques for anthropometric markers in each G2A study is provided in **Supplementary Methods**.

### AnthropoAge and AnthropoAgeAccel

AnthropoAge was previously developed and validated in the third and fourth National Health and Nutrition Examination Surveys (NHANES III and IV) by our team as a proxy of BA^17^. A detailed description on how AnthropoAge is calculated is available in **Supplementary Methods**. Briefly, AnthropoAge was created by using CA and anthropometry to predict 10-year mortality risk with sex-specific proportional hazards models following the Gompertz distribution. For this study, we employed the simplified version of AnthropoAge, which uses CA in years, BMI and WHtR. To measure biological age acceleration, we estimated AnthropoAgeAccel by removing the effect of CA using the residuals from a linear model regressing AnthropoAge onto CA^17,23^ and used this metric to define whether participants presented accelerated aging (see details in *Statistical analysis*).

### Classification of race and ethnicity

To address differences in anthropometry across populations, AnthropoAge uses race/ethnicity in the shape parameter of the Gompertz distribution. In HRS, the assessment of race and ethnicity adheres to the Office of Management and Budget Standards ^24,25^, based on this, we used a race/ethnicity classification where HRS participants were coded as either (Non-Hispanic) White, (Non-Hispanic) Black, Hispanic/Latino or Other (which includes Asian, Native American, or Pacific Islander); given the lack of consensus to define race and ethnicity across the world, the rest of surveys included in this study also follow this classification: participants from ELSA were coded as White, those from MHAS and CRELES as Hispanic/Latino, and those from CHARLS as Other (referring to Asian, and more specifically, Chinese participants).

### Functionality, general health status and comorbidities

Functional decline in older adults is closely related to the diagnosis of cognitive impairment and dementia^26–28^ and is associated with a higher mortality risk in this population^29^. Here, we used self-reported deficits in activities of daily living (ADL) and instrumental activities of daily living (IADL)^30^ to assess the functional status of participants. Although there are multiple validated clinical scales to appraise ADL and IADL^31^, we only included a few items to enable comparisons across G2A studies. For ADL deficits, participants were asked if they had some difficulty in performing any of the following: bathing, eating, transferring in and out of bed, using the toilet, and walking across the room. For IADL deficits, the same question was asked about: managing money, taking medication, shopping for groceries, and preparing hot meals. Based on this, we created scores ranging from 0-5 (ADL) and 0-4 (IADL) representing the number of activities in which participants presented any difficulty. In the case of CHARLS, “walking across the room” was unavailable and was substituted by “dressing” to facilitate comparisons using five ADL items (**Supplementary Material**). Participant’s self-reported general health status was evaluated across all cohorts by asking “Would you say your health is”: 1) excellent, 2) very good, 3) good, 4) fair, 5) poor. Additionally, several comorbidities were evaluated by self-report across all G2A cohorts, including hypertension, diabetes, cancer, chronic lung disease, myocardial infarction, stroke, and arthritis. The number of comorbidities was used to assess multimorbidity (≥2 comorbidities) in selected analyses.

### End-of-life data

For HRS, ELSA, MHAS and CHARLS, mortality was ascertained via a verbal autopsy by a family member proxy during an interview in a follow-up wave. For CRELES, all Costa Rican citizen participants were linked to the birth and death registries using their ID-card number. HRS and MHAS collected information on mortality status up to the year 2021, however, this information was only available up to 2020 for CHARLS, 2012 for ELSA and 2010 for CRELES (**Figure 1**). All studies collected the approximate date of death of deceased individuals, which was used along with date of baseline interview to estimate follow-up time in person-years (date of last interview was used for deceased individuals for whom date of death was not available). Individuals who had not died at the end of follow-up were censored.

**Figure 1.**
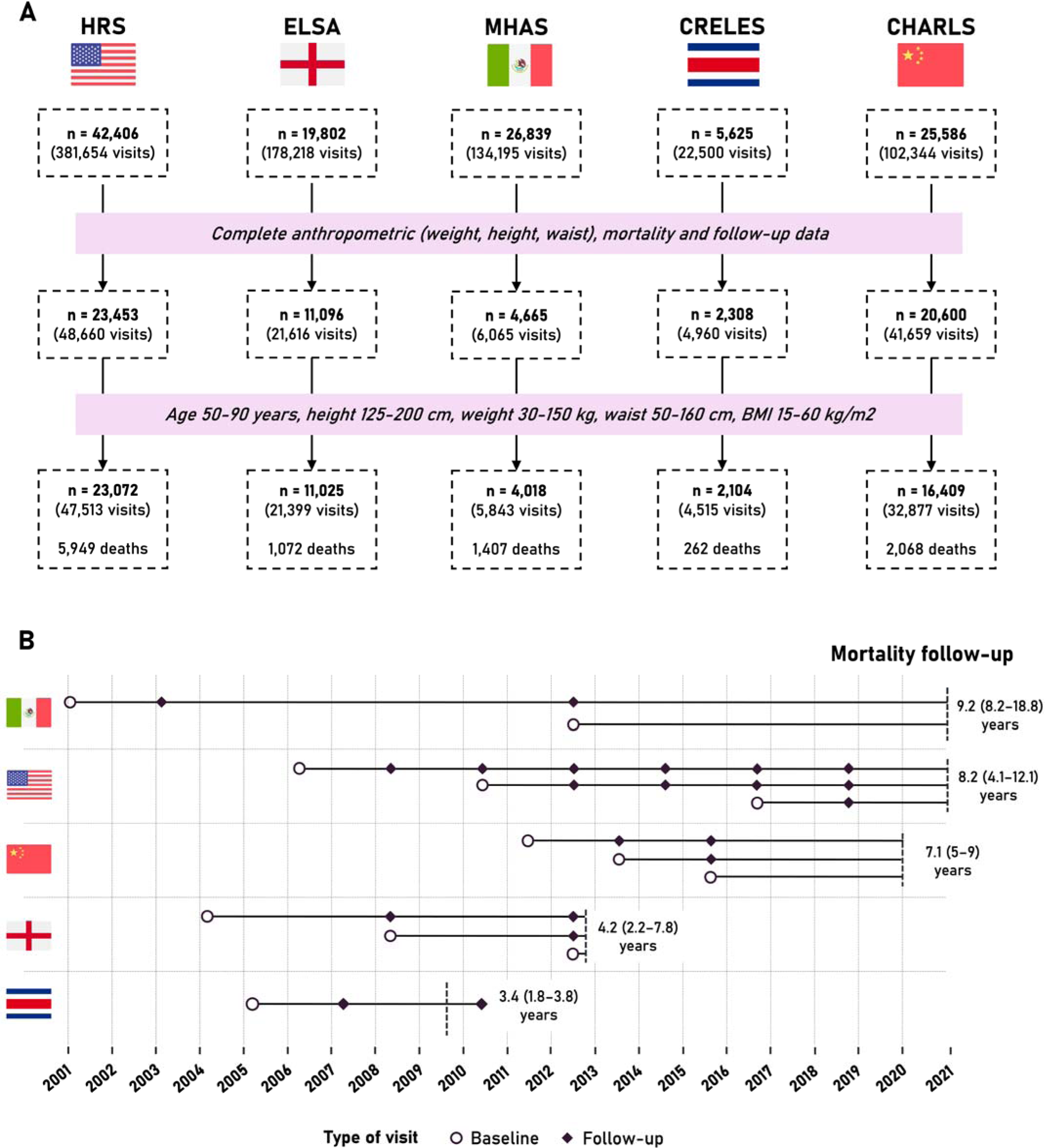
(**A**) Flowchart of participant selection for each individual survey from the harmonized Gateway to Global Aging (G2A) data. Participants were removed from our study if they had missing data on mortality, follow-up, or anthropometry, if they were aged <50 or >90 years old (to enhance comparability between surveys) or if they had anthropometric measurements outside of the ranges shown (to avoid unstable estimations). (**B**) Representation of follow-up among participants included in our study. All surveys (except CRELES) were comprised of multiple cohorts; circles represent the year of the first visit in each cohort and diamonds represent subsequent visits. The final sample size of the overall study was 56,628, with a total of 10,758 deaths over a joint median follow up of 7.6 years (IQR 3.9–9.1 years).

## Statistical Analysis

We restricted all analyses to individuals with complete mortality and anthropometric data, aged 50-90 years old, and with anthropometric measurements within the ranges observed in NHANES, where AnthropoAge was originally developed (height between 125-200cm, weight between 30-150kg, waist circumference between 50-160cm and BMI between 10- 60 kg/m^2^) (**Figure 1**).

### Estimation of AnthropoAgeAccel

To calculate AnthropoAgeAccel, a proxy of age acceleration, we regressed AnthropoAge onto CA and obtained the residuals using a mixed effects linear model, where we included a random intercept for ID to account for intra-individual dependence of longitudinal data and a second random intercept for G2A study to reduce potential clustering of the effect in each country. This was done separately for men and women to address sex-based differences in body composition and aging^32^. Accelerated aging was defined with AnthropoAgeAccel values >0, while non-accelerated or delayed aging was defined with values ≤0. We also used AnthropoAgeAccel quartiles to assess participants with more pronounced deviations from CA. All materials and code employed to facilitate estimation of AnthropoAge and AnthropoAgeAccel have been deployed into the *AnthropoAge* R package (https://github.com/oyaxbell/AnthropoAgeR).

### Assessment of AnthropoAge predictive performance

We validated the capability of baseline AnthropoAge to predict all-cause mortality using sex-stratified Cox proportional hazard regression models. Predictive performance was evaluated using Uno’s c-statistic (*survival* R package^33^), and time-dependent areas under the receiver operating characteristic curves (tAUC) (*riskRegression* R package^34^), both of which are based on the non-parametric approach of inverse-probability of censoring weights^35,36^. Uno’s c-statistic was used to assess the performance of AnthropoAge over the entire follow-up period overall and after stratifying by race/ethnicity, sex, age, BMI and WHtR; while tAUC was used to assess its performance at specific follow-up times up to 10 years for HRS and MHAS, and 4 years for ELSA, CRELES and CHARLS. Comparisons of c-statistics were conducted using non-parametric z-score tests (*survcomp* R package^37^). Results were computed for all the studies combined (overall G2A population) as well as for each individual G2A study separately.

### Accelerated aging and all-cause mortality risk

We estimated the hazard ratio (HR) with 95% confidence intervals (95% CI) for all-cause mortality for accelerated aging (AnthropoAgeAccel >0) and AnthropoAgeAccel quartiles using Cox proportional hazard regression models with standardized survey weights. These models were stratified by sex and race/ethnicity and sequentially adjusted for CA, number of comorbidities, education level (primary or less, secondary, tertiary), smoking (never, former smoker, <10 cigarettes/day, ≥10 cigarettes/day), and alcohol intake (never, less than weekly, less than daily, daily). This analysis was performed on baseline measurements, however, we also used Cox models clustered by ID to account for longitudinal measurements as a sensitivity analysis. Lastly, to visualize the additive effect of comorbidities and accelerated aging on mortality risk, we used Kaplan-Meier curves stratified by the presence of accelerated aging and multimorbidity (≥2 comorbidities).

### Changes in AnthropoAge and AnthropoAgeAccel over time

To evaluate trends of AnthropoAge as a function of time, we fitted weighted generalized estimating equation (GEE) models with a Gaussian variance function, where AnthropoAge was the dependent variable and years of follow-up was the predictor; robust sandwich standard errors and an autoregressive correlation structure (measures closer in time are more correlated) were used (*geepack* R package^38,39^). If this model had a β-coefficient >1, we considered that the rate of population aging occurred, on average, faster than expected with each year of follow-up, on the other hand, if the β-coefficient was <1 then the population aged, on average, slower than expected with each year of follow-up. We also conducted a sensitivity analysis stratifying these trends by sex for each study. To identify trends of age acceleration, we estimated the prevalence of accelerated aging and the distribution of AnthropoAgeAccel quartiles using survey weights.

### Association of AnthropoAgeAccel with ADL, IADL and comorbidities

We hypothesized that changes of AnthropoAgeAccel would also be predictive of functional decline, changes in self-reported health, and new onset of chronic age-related diseases independently of CA. To test this hypothesis, we fitted weighted GEE models with a Poisson variance function using robust sandwich standard errors and an autoregressive correlation structure, which we used to estimate rate ratios (RR) with 95% CI for AnthropoAgeAccel and accelerated aging, with the follow-up time in years (log-transformed) as the regression offset; all models were fully adjusted as specified above and considered clustering at an individual level. For all outcomes, we fitted a GEE model with only CA as a predictor and a second model adding AnthropoAgeAccel and compared the models using the quasi-likelihood information criterion (QIC), where negative values of the QIC difference between models (ΔQIC = QIC_M2_ – QIC_M1_) indicate that the addition of AnthropoAgeAccel improves the model’s fit. In the overall G2A analysis, these models were fitted in population at-risk, thus, we excluded participants who already had the outcome at baseline to identify new onset of functional deficits and comorbidities; however, when we stratified by country, we did not exclude these individuals to increase statistical power. All statistical analyses were conducted using R version 4.2.1 and p-value thresholds are estimated for a two-sided significance level of α=0.05.

## RESULTS

### Study population

Among a total of 120,258 participants across all five G2A studies, 62,122 had complete anthropometric data (weight, height, and waist circumference), mortality follow-up and survey weights. Of them, 55,628 (study population) were aged 50-90 years old and had anthropometric measurements within specified ranges, comprising a total of 117,147 longitudinal evaluations. A detailed flowchart of participant selection and follow-up evaluations is presented in **Figure 1** (this figure represents exclusively the waves and subsets where anthropometry was assessed). Participants had a median age of 61 years (IQR 55-70 years), most of them were female (46%), and the population was predominantly Non-Hispanic White (45%), followed by other race/ethnicities (Asian) (31%) and Hispanic/Latino participants (16%). On average, participants in CRELES were older (median age 74 [68-81] years), while participants in CHARLS were younger (59 [53-66] years). Median BMI was 26.7 kg/m^2^ (IQR 23.4-30.6) and median WHtR was 0.58 (IQR 0.53-0.64), with CHARLS participants being significantly leaner (median BMI 23.3 [20.9- 25.9] kg/m^2^, median WHtR 0.54 [0.50-0.59]). Accordingly, median AnthropoAge was 61 years (IQR 55-70) for the overall population, 63 (56-72) years for HRS, 62 (55-70) years for ELSA, 63 (56-71) years for MHAS, 76 (69-83) years for CRELES, and 57 (51-64) years for CHARLS participants. Overall, most participants had primary or less as their education level (50%), were never smokers (50%) and did not consume alcohol (47%); however, these percentages varied significantly across countries. A complete description of demographic and health characteristics of included participants is outlined in **Supplementary Table 1**.

### Performance of AnthropoAge vs. CA for all- cause mortality

In the overall G2A population, the total follow-up amounted to 400,347 person-years; the longest median follow-up was recorded for HRS at 9.2 years (IQR 8.2-18.8), while the shortest was registered for CRELES at 3.4 years (IQR 1.8-3.8). During this period, 10,758 all-cause deaths were recorded (19%), with higher mortality rates observed in MHAS (1,407 deaths, 35%) and HRS (5,949 deaths, 26%) (**Figure 1**). AnthropoAge was predictive of all-cause mortality (HR 2.68, 95%CI 2.61-2.75 per 1SD increase), with a predictive performance (Uno’s c-statistic 0.774, 95%CI 0.755-0.793) superior to CA (Uno’s c-statistic 0.760, 95%CI 0.735-0.786, p<0.001 for difference vs. AnthropoAge). When stratified by individual G2A studies, we observed a better prediction for AnthropoAge in ELSA, followed by HRS, CHARLS, MHAS, and CRELES; in all studies, except for MHAS, the c-statistic for AnthropoAge was significantly greater than the one for CA (**Figure 2A, Supplementary Table 2**). When stratifying by race/ethnicity, we found that, although numerically superior, AnthropoAge was statistically superior to CA only in White and Asian participants (**Figure 2B, Supplementary Table 2**). The predictive capacity of AnthropoAge was better in younger participants and without comorbidities and was broadly similar across sex and BMI/WHtR quintiles (**Supplementary Figure 1**). The tAUC for mortality prediction in the overall population increased over time for both AnthropoAge and CA (**Figure 2C**) despite a decrease at 8-10 years (which can be explained by differences in follow-up time across studies). Throughout the study period, AnthropoAge displayed a superior discriminatory capacity than CA, as shown by the difference between their tAUC’s being consistently greater than zero (**Figure 2D**). When disaggregated by study, AnthropoAge had a consistently superior tAUC for HRS, ELSA, CHARLS, and CRELES; however, this was not the case for MHAS (**Supplementary Figure 2**). To explore whether differences in sampling design across the two MHAS cohorts could affect results, c-statistic and tAUC for the MHAS 2001 sample and for the 2012 refreshment are shown separately in **Supplementary Figure 3**; where AnthropoAge displays a marginally better performance in MHAS 2012 compared to MHAS 2001.

**Figure 2.**
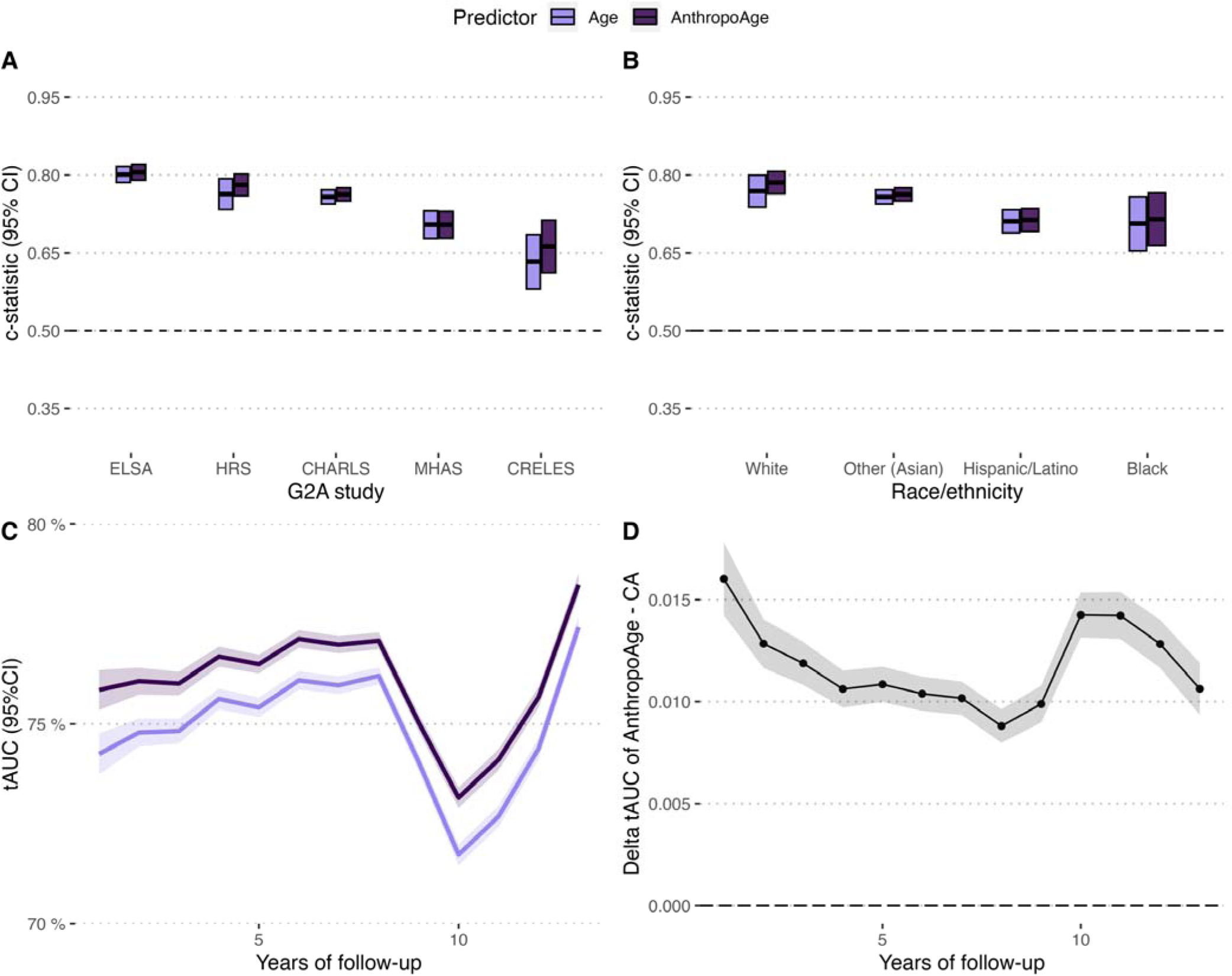
Predictive performance of AnthropoAge compared to Chronological Age (CA) as evaluated using Uno’s c-statistic for all-cause mortality stratified by individual Gateway to Global Aging dataset (**A**) and by race/ethnicity (**B**). The panels below show time-dependent performance of AnthropoAge compared to CA using time-dependent area under the receiving operating curve (tAUC) over 12 years of follow-up (**C**), as well as the delta tAUC for the same period comparing the performance of AnthropoAge minus the performance of CA (**D**).

### Accelerated aging and all-cause mortality

We sought to explore whether accelerated aging could independently predict the risk of all-cause mortality. In the overall G2A sample, participants with accelerated aging had an increased risk of death independently of CA, sex, race/ethnicity, number of comorbidities, education level, smoking and alcohol consumption compared to those without accelerated aging (HR 1.39 [95%CI 1.32-1.47]), and this result was replicated for HRS (1.41 [1.32- 1.51]), ELSA (1.27 [1.09-1.47]), MHAS (1.25 [1.03-1.53]), CRELES (1.59 [1.18-2.15]) and CHARLS (1.42 [1.26-1.61]) individually (**Figure 3A**). Furthermore, we observed that individuals with accelerated aging had a higher mortality risk than those without accelerated aging regardless of whether they had multimorbidity; in fact, we observed an additive effect where participants with both accelerated aging and multimorbidity presented the highest risk of death (**Figure 3B, Supplementary Figure 4**), and accelerated aging was still predictive of mortality in fully adjusted Cox models after stratifying by the presence of multimorbidity in the overall G2A population (**Supplementary Figure 5**). Lastly, when comparing the highest vs. the lowest AnthropoAgeAccel quartiles from longitudinal measurements, we observed an adjusted HR for all-cause mortality of 1.33 (95%CI 1.28-1.39) in the overall G2A population. This was replicated for HRS (1.40 [1.33- 1.47]), ELSA (1.45 [1.21-1.74]), CRELES (2.62 [1.83-3.75]) and CHARLS (1.28 [1.17-1.40]); although we did not observe a significant association for the overall MHAS population, this effect was present when examining the 2012 cohort separately (2.02 [1.18- 3.46]) (**Supplementary Table 3**).

**Figure 3.**
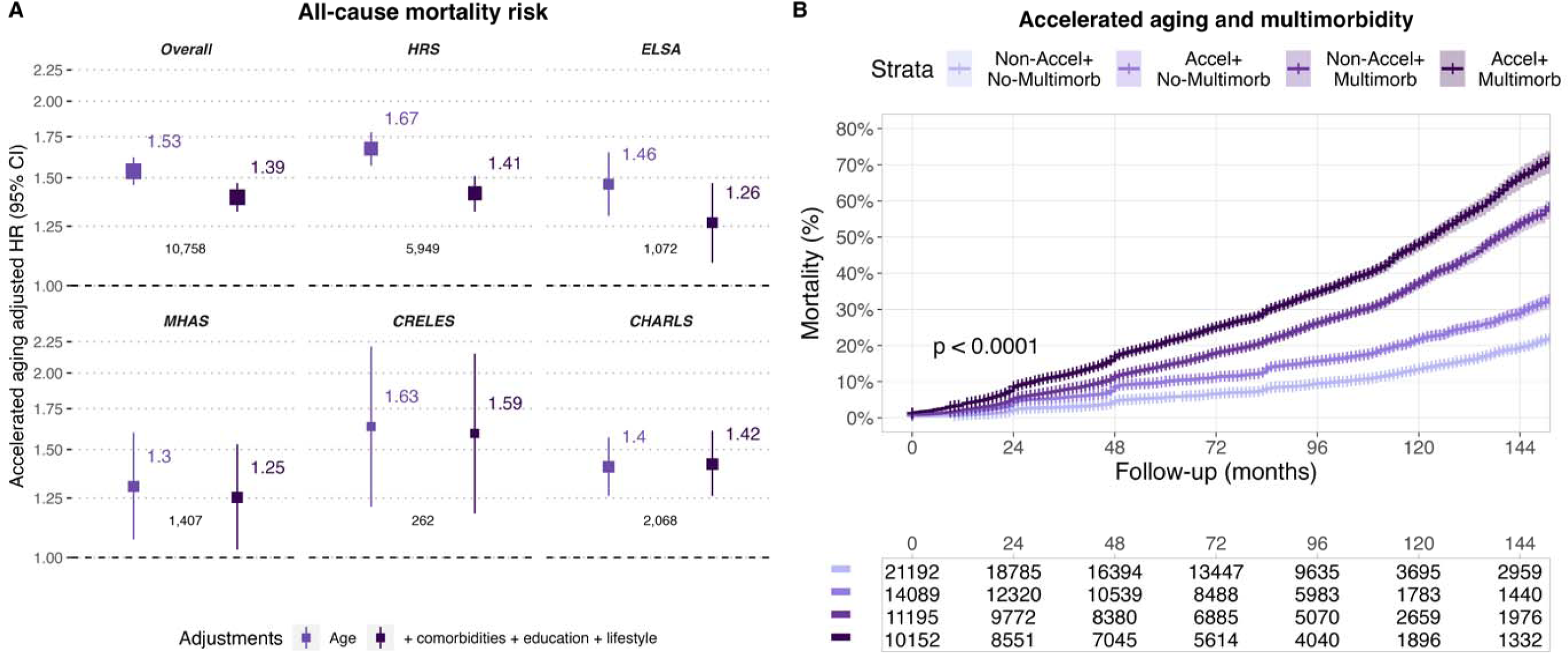
(**A**) Hazard ratio (95% CI) for all-cause mortality associated to accelerated aging in the overall G2A sample and stratified by survey. Estimates were obtained from Cox proportional hazard regression models adjusted 1) by chronological age and 2) by number of comorbidities, education level, smoking, and alcohol consumption. The larger top numbers represent the adjusted hazard ratio, and the smaller numbers represent the number of events in each group. (**B**) Kaplan-Meier curve comparing 12-year cumulative mortality risk stratified by the presence of accelerated aging (AnthropoAgeAccel >0) and multimorbidity (≥2 comorbidities) in individuals from the overall G2A sample; p-value was computed from a log-rank test for comparison of survival curves. Evaluated comorbidities include diabetes mellitus, hypertension, cancer, arthritis, stroke, myocardial infarction, and chronic obstructive pulmonary disease.

### AnthropoAge trends over time across G2A studies

Next, we used data from participants who were present at baseline and who had at least two visits to assess longitudinal changes in biological aging. Briefly, average AnthropoAge values increased at a faster rate than expected with each year of follow-up in HRS (β coefficient from linear GEE model: 1.07 [95% CI 1.06-1.08]), ELSA (1.08 [1.06-1.09]), CRELES (1.34, [1.29-1.40]), and CHARLS (1.02, [1.01-1.04]) (**Figure 4A**), while the slope of increase was also steep for MHAS, it was not significantly greater than 1. When exploring the prevalence of accelerated aging per year, we only observed a clear increasing trend for HRS and CRELES (**Figure 4B**). These trends were broadly similar for men and women in ELSA and CHARLS, however, men displayed a steeper increase in AnthropoAge with time in HRS (β for women 0.97 [0.94-1.00], β for men 1.08 [1.06-1.09]) and MHAS (β for women 1.00 [0.89-1.11], β for men 1.18 [1.07-1.29]), while women had a more pronounced change in CRELES (β for women 1.44 [1.36-1.52], β for men 1.19 [1.12- 1.26]) (**Supplementary Figures 6 and 7**). For all G2A studies, individuals with AnthropoAgeAccel values in the upper quartile sustained higher AnthropoAge values than CA, while those in the lower quartile generally remained below CA increases over time (**Supplementary Figure 8**).

**Figure 4.**
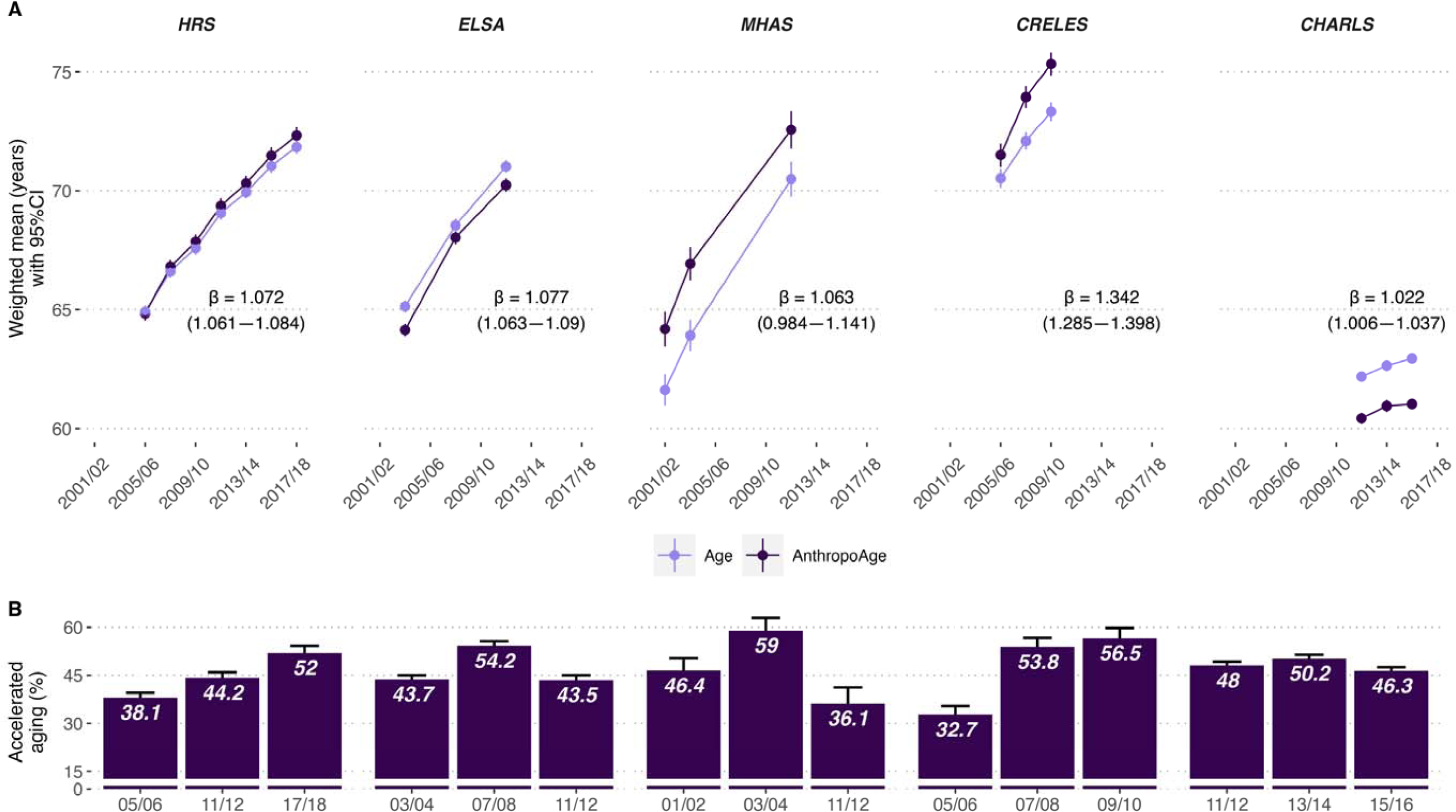
(**A**) Trends of weighted mean (95% CI) chronological age compared to weighted mean AnthropoAge plotted against follow-up time (biennial cycles) for each individual survey; β-coefficients were derived from generalized estimating equations with a Gaussian function (a β-coefficient >1 indicates that the rate of population aging occurs, on average, faster than expected with each year of follow-up). (**B**) Bar graphs represent weighted prevalence of accelerated aging (with 95% CI) defined as AnthropoAgeAccel >0 in biannual cycles for each survey. GEE models were performed in the total population study; however, to allow a better visualization of AnthropoAge changes through time, both plots only use information from participants who were present at baseline and who had at least two visits, yielding an overall sample size of 38,921 participants (HRS: 14,405. ELSA: 6,092. MHAS: 2,967. CRELES: 1,874. CHARLS: 13,605).

### Association of AnthropoAgeAccel with comorbidities, ADL or IADL deficits

We explored the ability of AnthropoAgeAccel to predict new-onset comorbidities, ADL/IADL deficits and changes in self-reported health in a longitudinal setting. Increasing AnthropoAgeAccel values (per 1-SD) predicted deterioration in self-reported health (RR 1.042 [95% CI 1.038-1.046]), new-onset ADL (1.23 [1.19-1.28]) and IADL deficits (1.24 [1.20-1.29]), as well as new-onset diabetes (1.10 [1.06-1.14]), hypertension (1.09 [1.06-1.12]), cancer (1.16 [1.12-1.21]), chronic lung disease (1.22 [1.16-1.27]), myocardial infarction (1.07 [1.04-1.11]), and stroke (1.13 [1.06-1.22]) independent of CA, sex, race/ethnicity, G2A cohort, education level, smoking and alcohol consumption; this was also observed for AnthropoAgeAccel >0. We also tested for AnthropoAgeAccel*Age interactions as a proxy of changes in AnthropoAgeAccel over time, however, this was only significantly associated with new-onset SRH and ADL deficit. For all outcomes, addition of AnthropoAgeAccel led to QIC minimization compared to models with CA alone, indicating improved model performance (**Table 1**). When disaggregating this analysis by G2A study, we observed similar trends for HRS, ELSA and CRELES; however, for CHARLS and MHAS AnthropoAgeAccel was only associated with ADL/IADL deficits (**Supplementary Figure 8**).

**TABLE 1.**
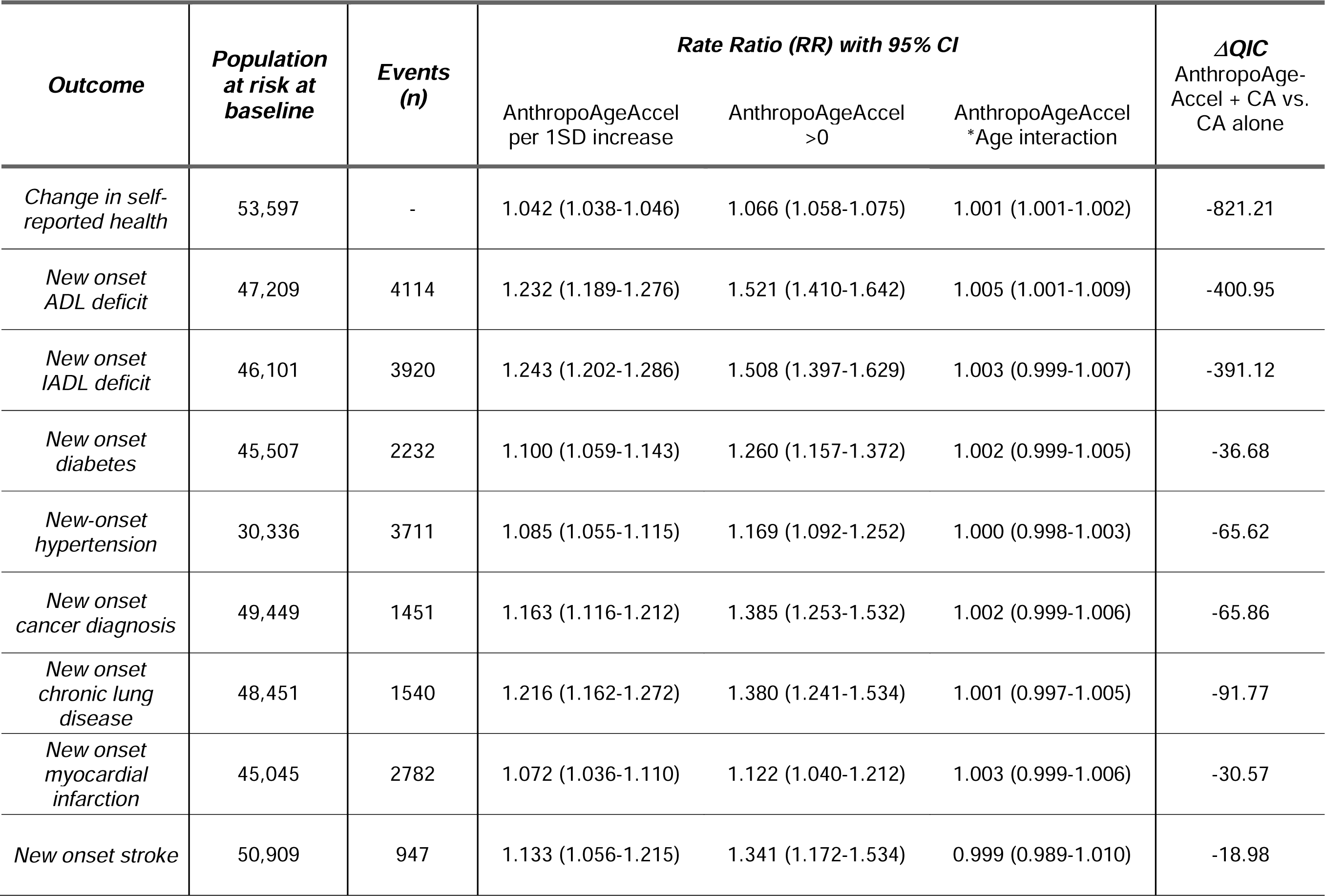
Performance of longitudinal evaluations of AnthropoAgeAcel and accelerated aging (AnthropoAgeAccel >0 years) for prediction of new onset of ADL/IADL deficits, change in self-reported health, or new self-reported diagnosis of age-related comorbidities evaluated in harmonized G2A data. Rate ratios (RR) with 95% CI were derived from weighted generalized estimating equations with a Poisson variance function using. All models were adjusted for chronological age (CA), sex, race/ethnicity, survey, education level, smoking and alcohol consumption and clustered at an individual level. Comparison of models with and without AnthropoAgeAccel were conducted using delta quasi-likelihood information criteria (ΔQIC), where negative values indicate that the addition of AnthropoAgeAccel improves the model’s goodness of fit.

| **Outcome** | **Population**<br>**at risk at**<br>**baseline** | **Events**<br>**(n)** | **Rate Ratio (RR) with 95% CI** |  |  | **ΔQIC**<br>**AnthropoAge-**<br>**Accel + CA vs.**<br>**CA alone** |
| --- | --- | --- | --- | --- | --- | --- |
|  |  |  | AnthropoAgeAccel<br>per 1SD increase | AnthropoAgeAccel<br>>0 | AnthropoAgeAccel<br>*Age interaction |  |
| *Change in self-*<br>*reported health* | 53,597 | - | 1.042 (1.038-1.046) | 1.066 (1.058-1.075) | 1.001 (1.001-1.002) | -821.21 |
| *New onset*<br>*ADL deficit* | 47,209 | 4114 | 1.232 (1.189-1.276) | 1.521 (1.410-1.642) | 1.005 (1.001-1.009) | -400.95 |
| *New onset*<br>*IADL deficit* | 46,101 | 3920 | 1.243 (1.202-1.286) | 1.508 (1.397-1.629) | 1.003 (0.999-1.007) | -391.12 |
| *New onset*<br>*diabetes* | 45,507 | 2232 | 1.100 (1.059-1.143) | 1.260 (1.157-1.372) | 1.002 (0.999-1.005) | -36.68 |
| *New-onset*<br>*hypertension* | 30,336 | 3711 | 1.085 (1.055-1.115) | 1.169 (1.092-1.252) | 1.000 (0.998-1.003) | -65.62 |
| *New onset*<br>*cancer diagnosis* | 49,449 | 1451 | 1.163 (1.116-1.212) | 1.385 (1.253-1.532) | 1.002 (0.999-1.006) | -65.86 |
| *New onset*<br>*chronic lung*<br>*disease* | 48,451 | 1540 | 1.216 (1.162-1.272) | 1.380 (1.241-1.534) | 1.001 (0.997-1.005) | -91.77 |
| *New onset*<br>*myocardial*<br>*infarction* | 45,045 | 2782 | 1.072 (1.036-1.110) | 1.122 (1.040-1.212) | 1.003 (0.999-1.006) | -30.57 |
| *New onset stroke* | 50,909 | 947 | 1.133 (1.056-1.215) | 1.341 (1.172-1.534) | 0.999 (0.989-1.010) | -18.98 |

## DISCUSSION

In this population-based, longitudinal, and cross-nationally harmonized study including 117,147 follow-up assessments of 55,628 participants 50-90 years from the U.S., England, Mexico, Costa Rica, and China, we examined the use of AnthropoAge as a proxy of BA across different populations. AnthropoAge showed a strong performance for prediction of all-cause mortality, with a discriminatory ability ranging from 70-80% that improved over time and remained superior to CA alone in the overall G2A population; however, it varied across studies and race/ethnicities and appears to have lower predictive value for Hispanic/Latino individuals. Additionally, using AnthropoAge as a tool to evaluate trends of biological aging across countries, we identified a faster aging rate for participants living in the United States, England, Costa Rica, and China, but no definitive trend for Mexican participants (only present in Mexican men). Furthermore, accelerated aging phenotypes derived using AnthropoAgeAccel were useful to predict a ∼39% risk increase in all-cause mortality, ∼6% higher rates of worsening of self-reported health, ∼50% higher rates of new-onset difficulties in ADL/IADL, and higher rates of new-onset age-related diseases independently of CA, sex, race/ethnicity, education level, lifestyle, and comorbidity profiles in all evaluated countries. Altogether, these results highlight the value of AnthropoAge as a potential longitudinal biomarker of aging and age-related diseases.

### Utility and challenges of BA markers

Aging is characterized by a progressive deterioration of multiple biological systems over time, which culminates in functional, cognitive, and physical decline, and ultimately leads to disability, dependence and comorbidity burden in aging individuals^40,41^. Despite the immense heterogeneity of this process, the implementation of BA measures into multidisciplinary clinical assessments of older adults is still limited^42^. In contrast, more widely recognized and implemented concepts to assess aging in a clinical setting are those of frailty (both the deficit accumulation and phenotype definitions) and multimorbidity (two or more coexisting diseases), often involve overt clinical manifestations whose progression may be halted but not necessarily reversed^7,43,44^, and manifest as a persistent decay of interacting physiologic systems, reduced functional and homeostatic reserve and decreased resilience^43^; nonetheless, the aging process in earlier stages of life may lead to deficits without overt clinical manifestations, which are often undetected by conventional aging assessments^45,46^, but may be captured by biomarkers that measure the pace of biological aging. Additionally, frailty and multimorbidity may be influenced by a wide range of molecular and sociodemographic determinants which often overlap with biological aging. This is why BA metrics may also predict frailty risk and increased comorbidity burden^7^ and reflect deterioration of biological systems at a stage where they could potentially be reversed and/or prevented. In our study, the predictive capacity of AnthropoAge for all-cause mortality was also observed in individuals without multimorbidity, supporting the view that this metric may detect nuanced changes in body-composition which underlie biological aging without overt clinical manifestations. Furthermore, the use of metrics of age acceleration such as AnthropoAgeAccel may aid to detect individuals at the highest risks of developing age-related outcomes. Specifically, we found that subjects in the highest AnthropoAgeAccel quartile in the United States, England, Costa Rica, and China had a 40%, 45%, 162%, and 28% higher risk of all-cause mortality, respectively, compared to subjects in the lowest AnthropoAgeAccel quartile (we did not observe this effect in the whole Mexican study, however, the risk increase was 102% when assessing the 2012 cohort). These findings strengthen the notion that BA metrics may be a useful, more nuanced alternative to conventional clinical assessments, which could help to detect early alterations which may benefit from preventive measures to promote healthier aging in the population^47,48^.

Recent aging research has focused on the development of longitudinal biomarkers of aging, particularly through the implementation of multi-omics (e.g., third generation epigenetic clocks). However, the large-scale implementation of these BA metrics in epidemiological studies has remain limited due to the costs and inaccessibility of such data; furthermore, given that aging is a highly heterogeneous process, multiple biomarkers of aging often evaluate distinct underlying processes^9,15^. Thus, BA metrics based on simple and accessible measurements may provide an easy overview of individual aging trajectories across different body systems^1,2^. Here, we show that changes in AnthropoAge and AnthropoAgeAccel are useful to detect mortality risk, declines in self-reported health and functionality, and new onset of age-related diseases over time, indicating that these BA metrics may be useful longitudinal biomarkers of aging. Furthermore, our results also represent a proof-of-concept of how the implementation of readily available aging biomarkers may be useful to track trends of biological aging at the population level, an approach which has gained interest to understand exposures which may influence age-related outcomes in diverse populations^49^. Although AnthropoAge was created as a second-generation aging biomarker to estimate 10-year all-cause mortality, previous research has shown that the usage of prospective data may lead second-generation aging biomarkers to better performance to detect age-related outcomes^50^, as demonstrated in our study.

### Effect of sex and race/ethnicity on body composition aging

Our previous evaluation of AnthropoAge in NHANES demonstrated that AnthropoAge captures a sexual dimorphism in body composition with aging, whereby accelerated aging in females is primarily associated with body-fat distribution (i.e., increased visceral fat depots and reduced subcutaneous adiposity) and in males it is associated with muscle mass decline and abdominal adiposity^17^, a difference that has been heavily attributed to changes over time in the effect of sex hormones on body composition, particularly in post- menopausal women^51^. Here, we observed that the predictive capacity of AnthropoAge and AnthropoAgeAccel was —although slightly superior in women— mostly sex-independent; however, further research should investigate whether longitudinal changes captured by AnthropoAge are associated with distinct body composition phenotypes and whether these indicate specific underlying biological processes which may intersect with other aging domains in different body systems.

Moreover, race and ethnicity have been widely shown to be important determinants of both body composition and aging^52^. When stratifying our analyses by country of origin, we found that changes in AnthropoAge and AnthropoAgeAccel may be useful to predict age-related outcomes in older adults from various ethnic and sociodemographic backgrounds, as results from the overall G2A population were broadly replicated in participants from HRS, ELSA, CRELES and CHARLS, although this was not the case for MHAS in most analyses. This could be a consequence of discrepancies in sampling design and in the selection of the anthropometric subset, as well as a reduced sample size and number of events. However, these results could also reflect unique aging patterns in Latin American populations, where social determinants of health, lifestyle, and a high burden of cardiometabolic disease can generate a larger impact on the population’s health^49^. In summary, implementation of AnthropoAge may be useful to evaluate trends of body-composition aging across different race/ethnicities, nevertheless, recalibration may be needed to improve its utility in underrepresented populations, particularly of Hispanic/Latino ethnicity, given the known differences in body composition and anthropometry across populations^32,53,54^.

## Strengths and limitations

Our study had several strengths including a population-based multi-national sample of older individuals which is representative at the national level, which allowed us to externally validate AnthropoAge across several race/ethnicities and socioeconomic backgrounds as a marker of mortality and age-related outcomes independently of common determinants of healthy aging. Second, by combining statistical methods developed to evaluate individual-level prediction with population-level average effects using longitudinal data, we were able to show that AnthropoAge may be a useful longitudinal aging biomarker and that its performance in selected populations (i.e., Hispanic/Latino individuals) may warrant further recalibrations to account for ethnic differences in the contribution of body composition to mortality risk. Third, by exploring an accessible aging biomarker which may be reproduced in large-scale epidemiological studies, we were able to show that AnthropoAge may be useful to detect changes in the rate of biological aging over time at the population level. However, we should also acknowledge some limitations which have to be considered to adequately frame the results of our work. By using harmonized datasets from the G2A platform and considering the effect of the study in all statistical analyses we were able to minimize differences between cohorts; however, some intrinsic differences in sampling and study design may influence interpretations of results (e.g., lower observed performance or non-association of relevant outcomes), furthermore, because definitions of exposures and outcomes are heterogeneous across different G2A studies, some variability was observed, namely difficulties in ADL’s and mortality ascertainment (verbal autopsy by proxy interview for all studies except CRELES, for which death registry linkage was used). Finally, because we implemented the simplified version of AnthropoAge, we may not be able to fully capture the spectrum of body composition aging reflected by the full measure with more diverse anthropometric measurements.

## Conclusion

In conclusion, AnthropoAge and AnthropoAgeAccel are useful proxies of BA and accelerated aging which capture mortality risk, functional decline, and risk of age-related diseases in diverse populations. We showed that longitudinal assessments of BA acceleration capture mortality risk, functional decline, and risk of chronic diseases independent of CA, comorbidities, education, and lifestyle, and that AnthropoAgeAccel detects individuals at a particularly high risk. Implementation of AnthropoAge in population- based longitudinal studies may allow for modeling trends of biological aging, permitting accessible assessment of faster or slower rates of aging at the population level for planning of public health interventions. Lastly, despite its strong overall and time-dependent performance for prediction of mortality, we observed heterogeneous results across different race/ethnicities, and recalibration of AnthropoAge may be warranted to improve its applicability in populations which are often underrepresented in aging studies, including Latin American, Southeast Asian, and African older adults.

## ACKNOWLEDGMENTS

This project was registered and approved by the Research Committee at Instituto Nacional de Geriatría, project number DI-PI-004-2024 and was supported by a grant provided by CONAHCyT, project number CBF2023-2024-467. CAFM, and JPE are enrolled at the PECEM Program of the Faculty of Medicine at UNAM. CAFM, JPE and DRG are supported by CONAHCyT. JAS was supported by Grant Number K23DK135798 from the NIH/NIDDK and by the Massachusetts General Hospital Executive Committee and Center for Diversity and Inclusion Physician-Scientist Development Award

## AUTHOR CONTRIBUTIONS

Research idea and study design: CAFM, OYBC; data acquisition: CAFM, DRG, JPE, OYBC; analysis/interpretation: CAFM, OYBC, DRG, JPE, NEAV; statistical analysis: CAFM, OYBC; manuscript drafting: CAFM, DRG, NEAV, JPE, DAR, CGP, LMGR, JAS, OYBC; supervision or mentorship: DAR, CGP, JAS, OYBC. Each author contributed important intellectual content during manuscript drafting or revision and accepts accountability for the overall work by ensuring that questions pertaining to the accuracy or integrity of any portion of the work are appropriately investigated and resolved.

## DATA AVAILABILITY

All code, datasets and materials are available for reproducibility of results at https://github.com/oyaxbell/g2a_anthropoage/

## SUPPLEMENTARY MATERIAL

### SUPPLEMENTARY METHODS

#### Overview of G2A longitudi3nal surveys

The following overview of the Gateway to Global Aging (G2A) surveys was obtained from the G2A Health and Retirement Studies overview page (https://g2aging.org/survey/overview), as well as from the individual cohort profiles of each study.

- Health and Retirement Study (HRS)^1^:

◦ Started in 1992 and is currently comprised of 15 biennial waves (last one in 2020).
◦ Nationally representative data of adults aged ≥51 years from private households in the U.S., with oversampling for African American and Hispanic households.
◦ Currently comprised of 8 cohorts (original HRS sample plus 7 refreshments), of which only 3 are included in our study due to availability of anthropometric data.
◦ Anthropometry is available from waves 8-14 every two waves among each half-sample of respondents (i.e., one half of participants has anthropometry for waves 8, 10, 12 and 14, while the other half has anthropometry for waves 9, 11 and 13).
- English Longitudinal Study of Ageing (ELSA)^2^:

◦ Started in 2002, comprised of 9 biennial waves (last in 2019); end of life interviews were only conducted up to wave 6 (2012).
◦ Nationally representative, adults from England aged ≥50 years, no oversampling.
◦ Comprised of 6 cohorts (original plus 5 refreshments), only 3 included in our study.
◦ Anthropometry is available in waves 2, 4 and 6 (2004, 2008 and 2012). It is also available for 50% of participants in wave 8 and for the remaining 50% of participants in wave 9; however, we did not include them in our study due to missing mortality data.
- Mexican Health and Aging Study (MHAS)^3^:

◦ Started in 2001, comprised of 5 waves (2001, 2003, 2012, 2015, 2019).
◦ Nationally representative at both rural and urban levels, adults from Mexico aged ≥50 years, oversampling (original cohort) in the six states that accounted for 40% of all migration to the USA (Durango, Guanajuato, Jalisco, Michoacán, Nayarit, Zacatecas) (https://enasem.org/Documentation/SurveyDesign_Esp.aspx).
◦ Comprised of 3 cohorts (original plus 2 refreshments), only 2 included in our study.
◦ Anthropometry available in a random subsample of participants from waves 1-2, and in the full sample of four states (one highly urbanized, one with high-migration, one relatively poor and one with a high prevalence of diabetes) from wave 3.
- Costa Rican Longevity and Healthy Aging Study (CRELES)^4^:

- Original CRELES pre-1945 cohort.

- Includes waves 1-3 (2004-2006, 2007-2008, 2009-2010).
- Nationally representative sample of Costa Rican adults born before 1945 (aged ≥60 years), with oversampling in participants aged ≥95 years old.
- All participants were eligible for anthropometric measurements.
- This is the cohort we included in our study.
- CRELES 1945-1955 retirement cohort.

- Includes waves 4-5 (2010-2011, 2012-2014).
- Nationally representative of Costa Rican adults born between 1945 and 1955 (aged 55 to 65 years old), with no oversampling.
- All participants were eligible for anthropometric measurements.
- This cohort was excluded from our study.
- China Health and Retirement Longitudinal Study (CHARLS)^5^:

◦ Started in 2011, comprised of 4 waves (2011, 2013, 2015, 2019).
◦ Nationally representative at both rural and urban levels, adults from China aged ≥45 years, with no oversampling.
◦ Comprised of 4 cohorts (original plus 3 refreshments), only 3 included in our study.
◦ All participants from waves 1-3 were eligible for anthropometric measurements.

#### Assessment of anthropometric variables across G2A studies

| Measurement |  | HRS | ELSA | MHAS | CRELES | CHARLS |
| --- | --- | --- | --- | --- | --- | --- |
| Weight | Availability | Waves 8-14 | Waves 2,4,6 | Waves 1-3 |  |  |
|  | Technique | Healthometer 830KL | Not detailed | Electronic portable scales | Life source scale | Unspecified scale |
| Height | Availability | Waves 8-14 | Waves 2,4,6 | Waves 1-3 |  |  |
|  | Technique | Tape measure | Stadiometer |  |  |  |
| Waist circumference | Availability | Waves 8-14 | Waves 2,4,6 | Waves 1-3 |  |  |
|  | Technique | Height of navel | Not detailed | Midway between iliac crest and costal margin |  | Height of navel |

##### AnthropoAge calculation

AnthropoAge was developed to predict 10-year all-cause mortality based on the work by Levine et al.^6^ using two parametric proportional hazards models following the Gompertz distribution:

1. Using only chronological age (CA) as the predictor.
2. Using CA and anthropometric measurements (weight, height, and waist in the simplified version).

Both models were stratified by sex, and the second model included race/ethnicity (Non-Hispanic White, Non-Hispanic Black, Hispanic/Latino, Other) in the shape parameter of the distribution.

In order to obtain an estimate of biological age (BA) in age units, we assumed the cumulative distribution functions (CDF’s) of each model to be approximately equal, implying:

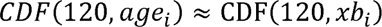

Where *age*_i_ is the linear combination of coefficients from the Gompertz model including only CA, and *xb*_i_ the linear combination coefficients from the model including both CA and anthropometric measurements. The solution to this equation is the i-th individual’s AnthropoAge, which is implied to be a function of the CDF at 120 months for the linear combination of anthropometric measurements (i.e., the risk of dying within the next 10 years predicted by CA and anthropometric measurements), expressed in units of age.

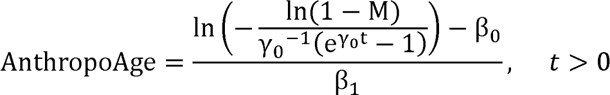

Where:

| Sex | $\gamma_0$ | $\beta_0$ | $\beta_1$ |
| --- | --- | --- | --- |
| Men | 0.00585 | -11.28580 | 0.07811 |
| Women | 0.00736 | -12.39100 | 0.08600 |

| Sex | Shape $\gamma_j$ | $xb$ | |
| --- | --- | --- | --- |
|  |  | Parameter | Coefficient |
| Men | NH White: 0.00605<br>NH Black: +0.00098<br>Hispanic: -0.00009<br>Other: -0.00440 | Rate | -19.0818 |
|  |  | CA | 0.0733 |
|  |  | BMI (OP,1) | -26.6759 |
|  |  | BMI (OP,2) | 12.3235 |
|  |  | WHtR | 9.7851 |
| Women | NH White: 0.00772<br>NH Black: +0.00039<br>Hispanic: -0.00079<br>Other: -0.00255 | Rate | -19.2580 |
|  |  | CA | 0.0818 |
|  |  | BMI (OP,1) | -20.8035 |
|  |  | BMI (OP,2) | 9.2458 |

#### Assessment of functionality across G2A studies

Activities of Daily Living (ADL) and Instrumental Activities of Daily Living (IADL) are commonly used to evaluate functional status and independence in multidimensional geriatric assessments^7^. ADLs comprise fundamental tasks regarding self-care and well-being (i.e., being able to bathe, eat, and use the bathroom), while IADLs are more complex tasks that involve executive functions and are essential to sustain independent life within home and in the community^8^. Although there are several validated clinical scores such as the Katz Index^9^ or the Lawton and Brody scale^10^ to assess ADL and IADL, only the following items were available across the G2A cohorts used in this study:

| **Metric** | **Activity** | **HRS** | **ELSA** | **MHAS** | **CRELES** | **CHARLS** |
| --- | --- | --- | --- | --- | --- | --- |
| **ADL** | **Bathing** | Waves 1-15 | Waves 1-9 | Waves 1-4 | Waves 1-5 | Waves 1-4 |
|  | **Eating** | Waves 1-15 | Waves 1-9 | Waves 1-4 | Waves 1-5 | Waves 1-4 |
|  | **Transferring in/out of bed** | Waves 1-15 | Waves 1-9 | Waves 1-4 | Waves 1-5 | Waves 1-4 |
|  | **Using toilet** | Waves 2-15 | Waves 1-9 | Waves 1-4 | Waves 1-5 | Waves 1-4 |
|  | **Walking across a room** | Waves 1-15 | Waves 1-9 | Waves 1-4 | Waves 1-5 | - |
|  | **Dressing** | Waves 1-15 | Waves 1-9 | Waves 1-4<br>(not in proxy interviews) | - | Waves 1-4 |
|  | **Sphincter control** | - | - | - | - | Waves 1-4 |
| **IADL** | **Managing money** | Waves 2-15 | Waves 1-9 | Waves 1-5 | Waves 1-5 | Waves 1-4 |
|  | **Taking medication** | Waves 2-15 | Waves 1-9 | Waves 1-5 | Waves 1-5 | Waves 1-4 |
|  | **Shopping for groceries** | Waves 2-15 | Waves 1-9 | Waves 1-5 | Waves 1-5 | Waves 1-4 |
|  | **Preparing hot meals** | Waves 2-15 | Waves 1-9 | Waves 1-5 | Waves 1-5 | Waves 1-4 |
|  | Using a telephone | Waves 2-15 | Waves 1-9 | - | - | Waves 2-4 |
|  | Using a map | Waves 1-15 | Waves 1-9 | - | - | - |

These variables were coded 0 if participants had no difficulty and 1 if they reported some difficulty in each activity. Based on this, we constructed scores to assess ADL and IADL by summing the variables that were available across all five studies. Scores were constructed in participants in whom at least one item was not missing, as recommended in the G2A Harmonized Codebooks (available from https://g2aging.org/hrd/get-data):

- ***ADL score*** (range: 0-5) = Bathing + Eating + Transferring in/out of bed + Using the toilet + Walking across the room.

◦ For CHARLS, we used dressing instead of walking across the room.
- ***IADL score*** (range: 0-4) = Managing money + Taking medications + Shopping for groceries + Preparing hot meals.

#### Ethical disclosures for G2A studies

- HRS was approved by the Health Sciences/Behavioral Sciences Institutional Review Board at the University of Michigan.
- ELSA was approved by the National Health Service Research Ethics Committees under the National Research and Ethics Service.
- MHAS was approved by the Institutional Review Board or Ethics Committee of the University of Texas Medical Branch, the National Institute of Statistics and Geography, and the National Institute of Public Health.
- CRELES was approved by the Ethical Science Committee of the University of Costa Rica.
- was approved by the Biomedical Ethics Review Committee of Peking University.

## SUPPLEMENTARY TABLES

**Supplementary Table 1.**
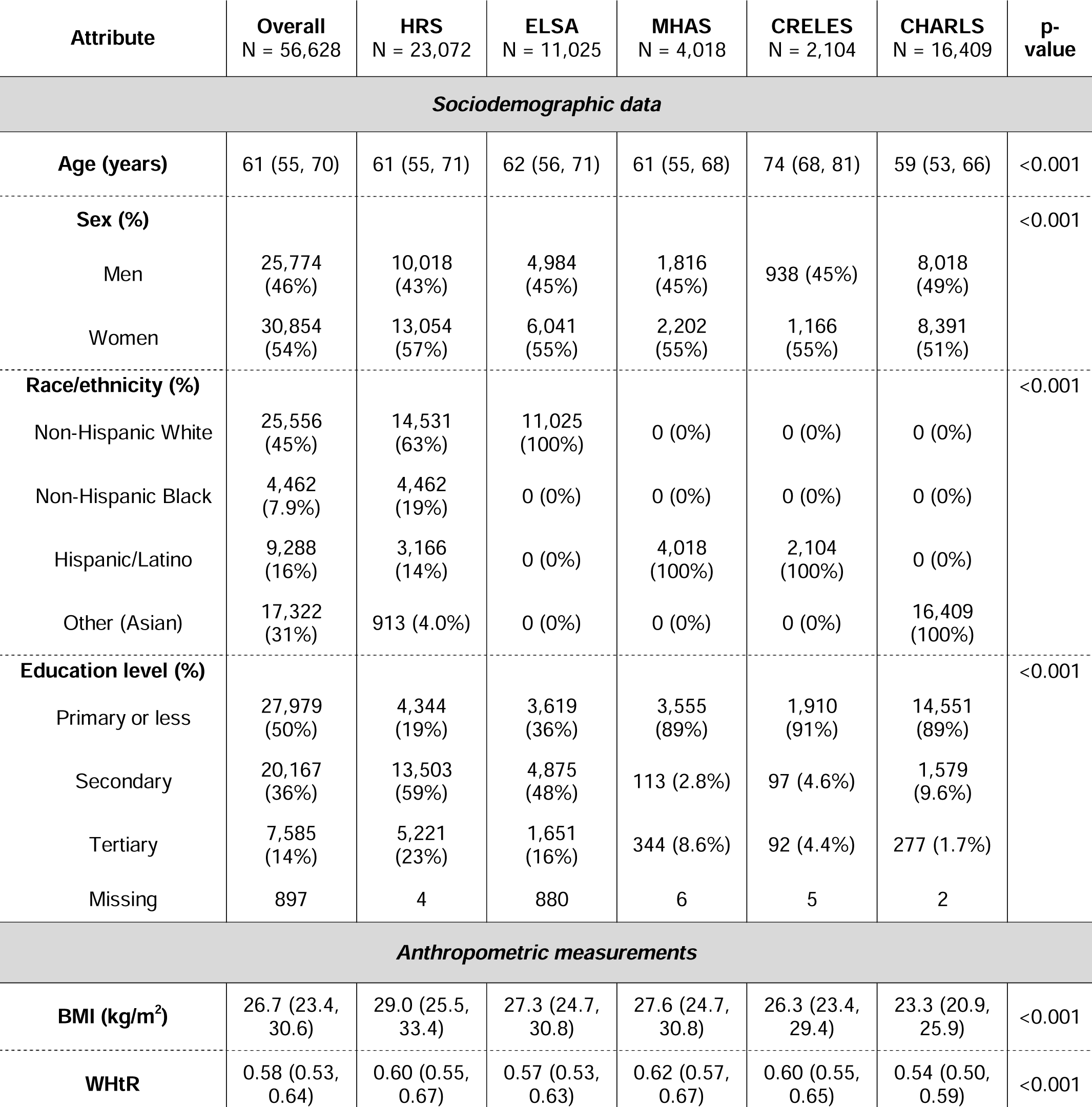
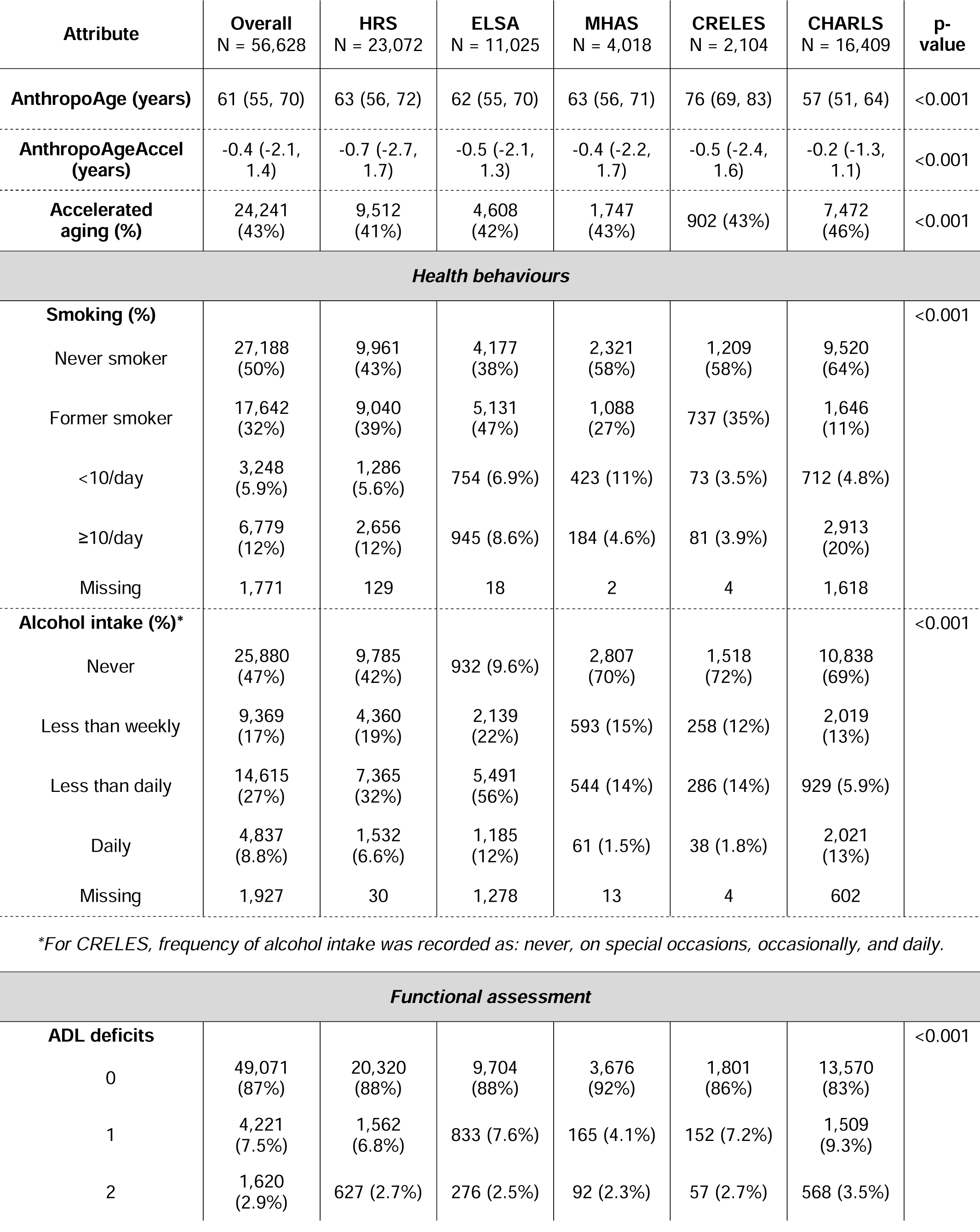
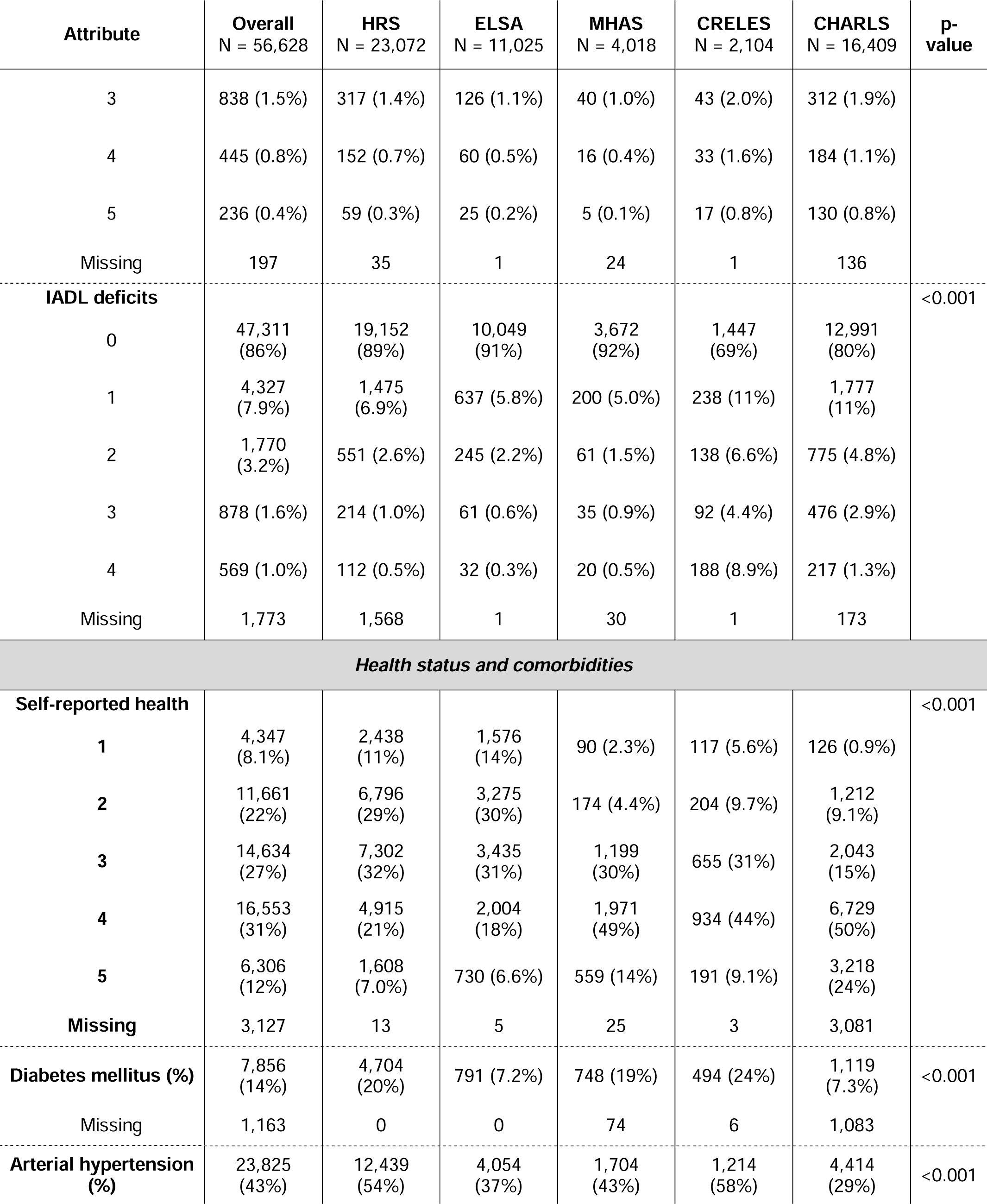
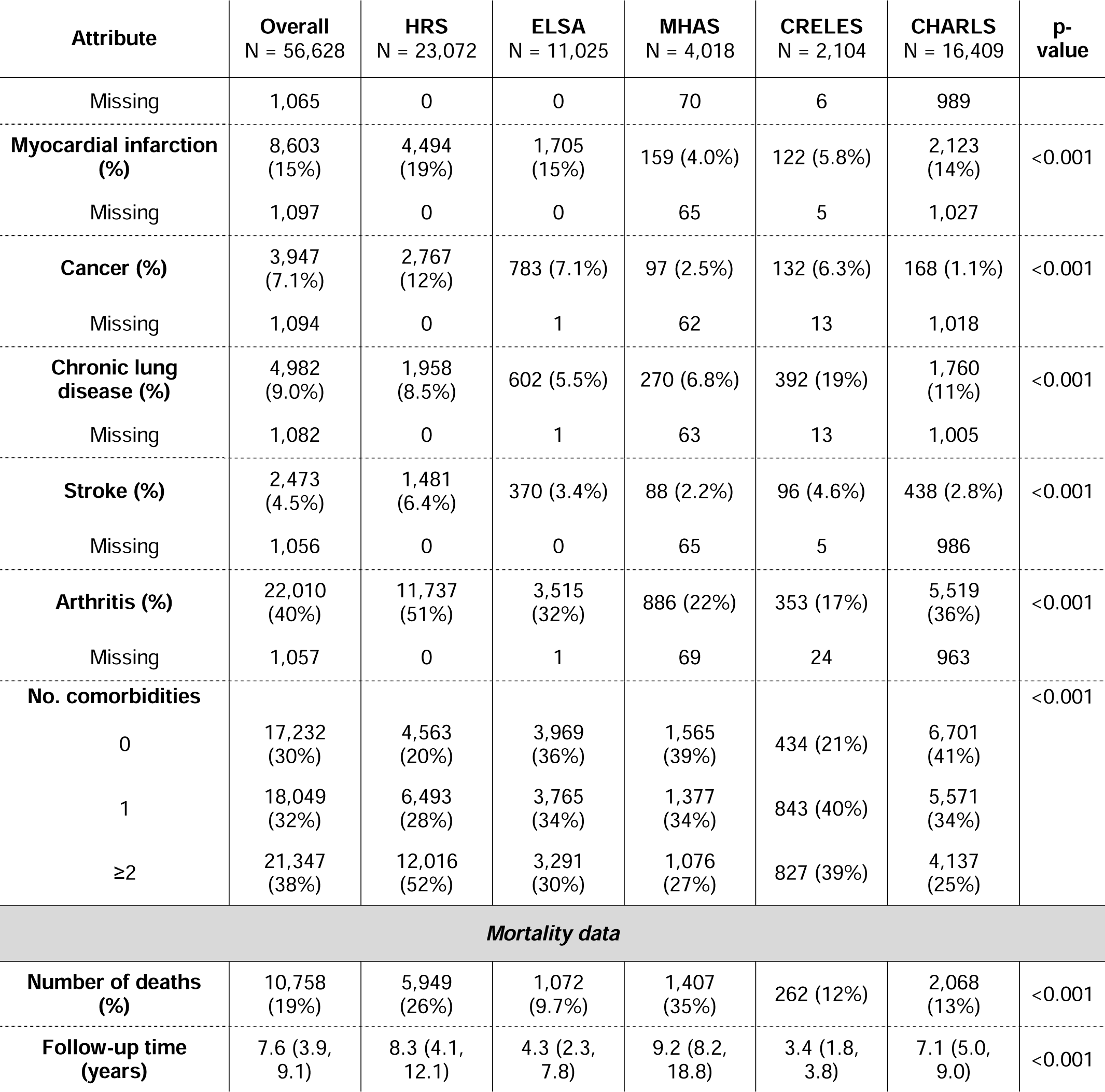
Baseline sociodemographic characteristics, anthropometric measurements, functional assessment, health status, and mortality data of participants aged 50-90 years (overall and stratified by cohort). Continuous variables are presented as median (with interquartile range), and categorical variables as absolute frequency (with percentage). P-values are computed using Kruskal- Walli’s rank sum test (continuous variables) or Pearson’s Chi-squared test (categorical variables).

| Attribute | Overall<br>N = 56,628 | HRS<br>N = 23,072 | ELSA<br>N = 11,025 | MHAS<br>N = 4,018 | CRELES<br>N = 2,104 | CHARLS<br>N = 16,409 | p-value |
| --- | --- | --- | --- | --- | --- | --- | --- |
| **Sociodemographic data** |  |  |  |  |  |  |  |
| **Age (years)** | 61 (55, 70) | 61 (55, 71) | 62 (56, 71) | 61 (55, 68) | 74 (68, 81) | 59 (53, 66) | <0.001 |
| **Sex (%)** |  |  |  |  |  |  | <0.001 |
| Men | 25,774<br>(46%) | 10,018<br>(43%) | 4,984<br>(45%) | 1,816<br>(45%) | 938 (45%) | 8,018<br>(49%) |  |
| Women | 30,854<br>(54%) | 13,054<br>(57%) | 6,041<br>(55%) | 2,202<br>(55%) | 1,166<br>(55%) | 8,391<br>(51%) |  |
| **Race/ethnicity (%)** |  |  |  |  |  |  | <0.001 |
| Non-Hispanic White | 25,556<br>(45%) | 14,531<br>(63%) | 11,025<br>(100%) | 0 (0%) | 0 (0%) | 0 (0%) |  |
| Non-Hispanic Black | 4,462<br>(7.9%) | 4,462<br>(19%) | 0 (0%) | 0 (0%) | 0 (0%) | 0 (0%) |  |
| Hispanic/Latino | 9,288<br>(16%) | 3,166<br>(14%) | 0 (0%) | 4,018<br>(100%) | 2,104<br>(100%) | 0 (0%) |  |
| Other (Asian) | 17,322<br>(31%) | 913 (4.0%) | 0 (0%) | 0 (0%) | 0 (0%) | 16,409<br>(100%) |  |
| **Education level (%)** |  |  |  |  |  |  | <0.001 |
| Primary or less | 27,979<br>(50%) | 4,344<br>(19%) | 3,619<br>(36%) | 3,555<br>(89%) | 1,910<br>(91%) | 14,551<br>(89%) |  |
| Secondary | 20,167<br>(36%) | 13,503<br>(59%) | 4,875<br>(48%) | 113 (2.8%) | 97 (4.6%) | 1,579<br>(9.6%) |  |
| Tertiary | 7,585<br>(14%) | 5,221<br>(23%) | 1,651<br>(16%) | 344 (8.6%) | 92 (4.4%) | 277 (1.7%) |  |
| Missing | 897 | 4 | 880 | 6 | 5 | 2 |  |
| **Anthropometric measurements** |  |  |  |  |  |  |  |
| **BMI (kg/m<sup>2</sup>)** | 26.7 (23.4,<br>30.6) | 29.0 (25.5,<br>33.4) | 27.3 (24.7,<br>30.8) | 27.6 (24.7,<br>30.8) | 26.3 (23.4,<br>29.4) | 23.3 (20.9,<br>25.9) | <0.001 |
| **WHtR** | 0.58 (0.53,<br>0.64) | 0.60 (0.55,<br>0.67) | 0.57 (0.53,<br>0.63) | 0.62 (0.57,<br>0.67) | 0.60 (0.55,<br>0.65) | 0.54 (0.50,<br>0.59) | <0.001 |

| Attribute | Overall<br>N = 56,628 | HRS<br>N = 23,072 | ELSA<br>N = 11,025 | MHAS<br>N = 4,018 | CRELES<br>N = 2,104 | CHARLS<br>N = 16,409 | p-value |
| --- | --- | --- | --- | --- | --- | --- | --- |
| **AnthroAge (years)** | 61 (55, 70) | 63 (56, 72) | 62 (55, 70) | 63 (56, 71) | 76 (69, 83) | 57 (51, 64) | <0.001 |
| **AnthroAgeAccel**<br>**(years)** | -0.4 (-2.1,<br>1.4) | -0.7 (-2.7,<br>1.7) | -0.5 (-2.1,<br>1.3) | -0.4 (-2.2,<br>1.7) | -0.5 (-2.4,<br>1.6) | -0.2 (-1.3,<br>1.1) | <0.001 |
| **Accelerated**<br>**aging (%)** | 24,241<br>(43%) | 9,512<br>(41%) | 4,608<br>(42%) | 1,747<br>(43%) | 902 (43%) | 7,472<br>(46%) | <0.001 |
| **Health behaviours** |  |  |  |  |  |  |  |
| **Smoking (%)** |  |  |  |  |  |  | <0.001 |
| Never smoker | 27,188<br>(50%) | 9,961<br>(43%) | 4,177<br>(38%) | 2,321<br>(58%) | 1,209<br>(58%) | 9,520<br>(64%) |  |
| Former smoker | 17,642<br>(32%) | 9,040<br>(39%) | 5,131<br>(47%) | 1,088<br>(27%) | 737 (35%) | 1,646<br>(11%) |  |
| <10/day | 3,248<br>(5.9%) | 1,286<br>(5.6%) | 754 (6.9%) | 423 (11%) | 73 (3.5%) | 712 (4.8%) |  |
| ≥10/day | 6,779<br>(12%) | 2,656<br>(12%) | 945 (8.6%) | 184 (4.6%) | 81 (3.9%) | 2,913<br>(20%) |  |
| Missing | 1,771 | 129 | 18 | 2 | 4 | 1,618 |  |
| **Alcohol intake (%)\*** |  |  |  |  |  |  | <0.001 |
| Never | 25,880<br>(47%) | 9,785<br>(42%) | 932 (9.6%) | 2,807<br>(70%) | 1,518<br>(72%) | 10,838<br>(69%) |  |
| Less than weekly | 9,369<br>(17%) | 4,360<br>(19%) | 2,139<br>(22%) | 593 (15%) | 258 (12%) | 2,019<br>(13%) |  |
| Less than daily | 14,615<br>(27%) | 7,365<br>(32%) | 5,491<br>(56%) | 544 (14%) | 286 (14%) | 929 (5.9%) |  |
| Daily | 4,837<br>(8.8%) | 1,532<br>(6.6%) | 1,185<br>(12%) | 61 (1.5%) | 38 (1.8%) | 2,021<br>(13%) |  |
| Missing | 1,927 | 30 | 1,278 | 13 | 4 | 602 |  |

\*For CRELES, frequency of alcohol intake was recorded as: never, on special occasions, occasionally, and daily.

|  |  |  |  |  |  |  |  |
| --- | --- | --- | --- | --- | --- | --- | --- |
| **Functional assessment** |  |  |  |  |  |  |  |
| **ADL deficits** |  |  |  |  |  |  | <0.001 |
| 0 | 49,071<br>(87%) | 20,320<br>(88%) | 9,704<br>(88%) | 3,676<br>(92%) | 1,801<br>(86%) | 13,570<br>(83%) |  |
| 1 | 4,221<br>(7.5%) | 1,562<br>(6.8%) | 833 (7.6%) | 165 (4.1%) | 152 (7.2%) | 1,509<br>(9.3%) |  |
| 2 | 1,620<br>(2.9%) | 627 (2.7%) | 276 (2.5%) | 92 (2.3%) | 57 (2.7%) | 568 (3.5%) |  |

| Attribute | Overall<br>N = 56,628 | HRS<br>N = 23,072 | ELSA<br>N = 11,025 | MHAS<br>N = 4,018 | CRELES<br>N = 2,104 | CHARLS<br>N = 16,409 | p-value |
| --- | --- | --- | --- | --- | --- | --- | --- |
| 3 | 838 (1.5%) | 317 (1.4%) | 126 (1.1%) | 40 (1.0%) | 43 (2.0%) | 312 (1.9%) |  |
| 4 | 445 (0.8%) | 152 (0.7%) | 60 (0.5%) | 16 (0.4%) | 33 (1.6%) | 184 (1.1%) |  |
| 5 | 236 (0.4%) | 59 (0.3%) | 25 (0.2%) | 5 (0.1%) | 17 (0.8%) | 130 (0.8%) |  |
| Missing | 197 | 35 | 1 | 24 | 1 | 136 |  |
| **IADL deficits** |  |  |  |  |  |  | <0.001 |
| 0 | 47,311<br>(86%) | 19,152<br>(89%) | 10,049<br>(91%) | 3,672<br>(92%) | 1,447<br>(69%) | 12,991<br>(80%) |  |
| 1 | 4,327<br>(7.9%) | 1,475<br>(6.9%) | 637 (5.8%) | 200 (5.0%) | 238 (11%) | 1,777<br>(11%) |  |
| 2 | 1,770<br>(3.2%) | 551 (2.6%) | 245 (2.2%) | 61 (1.5%) | 138 (6.6%) | 775 (4.8%) |  |
| 3 | 878 (1.6%) | 214 (1.0%) | 61 (0.6%) | 35 (0.9%) | 92 (4.4%) | 476 (2.9%) |  |
| 4 | 569 (1.0%) | 112 (0.5%) | 32 (0.3%) | 20 (0.5%) | 188 (8.9%) | 217 (1.3%) |  |
| Missing | 1,773 | 1,568 | 1 | 30 | 1 | 173 |  |
| **Health status and comorbidities** |  |  |  |  |  |  |  |
| **Self-reported health** |  |  |  |  |  |  | <0.001 |
| 1 | 4,347<br>(8.1%) | 2,438<br>(11%) | 1,576<br>(14%) | 90 (2.3%) | 117 (5.6%) | 126 (0.9%) |  |
| 2 | 11,661<br>(22%) | 6,796<br>(29%) | 3,275<br>(30%) | 174 (4.4%) | 204 (9.7%) | 1,212<br>(9.1%) |  |
| 3 | 14,634<br>(27%) | 7,302<br>(32%) | 3,435<br>(31%) | 1,199<br>(30%) | 655 (31%) | 2,043<br>(15%) |  |
| 4 | 16,553<br>(31%) | 4,915<br>(21%) | 2,004<br>(18%) | 1,971<br>(49%) | 934 (44%) | 6,729<br>(50%) |  |
| 5 | 6,306<br>(12%) | 1,608<br>(7.0%) | 730 (6.6%) | 559 (14%) | 191 (9.1%) | 3,218<br>(24%) |  |
| Missing | 3,127 | 13 | 5 | 25 | 3 | 3,081 |  |
| **Diabetes mellitus (%)** | 7,856<br>(14%) | 4,704<br>(20%) | 791 (7.2%) | 748 (19%) | 494 (24%) | 1,119<br>(7.3%) | <0.001 |
| Missing | 1,163 | 0 | 0 | 74 | 6 | 1,083 |  |
| **Arterial hypertension (%)** | 23,825<br>(43%) | 12,439<br>(54%) | 4,054<br>(37%) | 1,704<br>(43%) | 1,214<br>(58%) | 4,414<br>(29%) | <0.001 |

| **Attribute** | **Overall**<br>**N = 56,628** | **HRS**<br>**N = 23,072** | **ELSA**<br>**N = 11,025** | **MHAS**<br>**N = 4,018** | **CRELES**<br>**N = 2,104** | **CHARLS**<br>**N = 16,409** | **p-**<br>**value** |
| --- | --- | --- | --- | --- | --- | --- | --- |
| Missing | 1,065 | 0 | 0 | 70 | 6 | 989 |  |
| **Myocardial infarction (%)** | 8,603 (15%) | 4,494 (19%) | 1,705 (15%) | 159 (4.0%) | 122 (5.8%) | 2,123 (14%) | <0.001 |
| Missing | 1,097 | 0 | 0 | 65 | 5 | 1,027 |  |
| **Cancer (%)** | 3,947 (7.1%) | 2,767 (12%) | 783 (7.1%) | 97 (2.5%) | 132 (6.3%) | 168 (1.1%) | <0.001 |
| Missing | 1,094 | 0 | 1 | 62 | 13 | 1,018 |  |
| **Chronic lung disease (%)** | 4,982 (9.0%) | 1,958 (8.5%) | 602 (5.5%) | 270 (6.8%) | 392 (19%) | 1,760 (11%) | <0.001 |
| Missing | 1,082 | 0 | 1 | 63 | 13 | 1,005 |  |
| **Stroke (%)** | 2,473 (4.5%) | 1,481 (6.4%) | 370 (3.4%) | 88 (2.2%) | 96 (4.6%) | 438 (2.8%) | <0.001 |
| Missing | 1,056 | 0 | 0 | 65 | 5 | 986 |  |
| **Arthritis (%)** | 22,010 (40%) | 11,737 (51%) | 3,515 (32%) | 886 (22%) | 353 (17%) | 5,519 (36%) | <0.001 |
| Missing | 1,057 | 0 | 1 | 69 | 24 | 963 |  |
| **No. comorbidities** |  |  |  |  |  |  | <0.001 |
| 0 | 17,232 (30%) | 4,563 (20%) | 3,969 (36%) | 1,565 (39%) | 434 (21%) | 6,701 (41%) |  |
| 1 | 18,049 (32%) | 6,493 (28%) | 3,765 (34%) | 1,377 (34%) | 843 (40%) | 5,571 (34%) |  |
| ≥2 | 21,347 (38%) | 12,016 (52%) | 3,291 (30%) | 1,076 (27%) | 827 (39%) | 4,137 (25%) |  |
| **Mortality data** |  |  |  |  |  |  |  |
| **Number of deaths (%)** | 10,758 (19%) | 5,949 (26%) | 1,072 (9.7%) | 1,407 (35%) | 262 (12%) | 2,068 (13%) | <0.001 |
| **Follow-up time (years)** | 7.6 (3.9, 9.1) | 8.3 (4.1, 12.1) | 4.3 (2.3, 7.8) | 9.2 (8.2, 18.8) | 3.4 (1.8, 3.8) | 7.1 (5.0, 9.0) | <0.001 |

**Supplementary Table 2.** Comparison of Harrel’s c-statistic using z-scores (AnthropoAge vs CA) for prediction of all-cause mortality in the overall population, and stratifying by G2A survey, race/ethnicity, sex, and number of comorbidities.

| Strata | AnthroAge<br>c-statistic (95% CI) | Age<br>c-statistic (95% CI) | Difference | P-value |
| --- | --- | --- | --- | --- |
| **By G2A study** |  |  |  |  |
| **Overall** | 0.772 (0.765-0.779) | 0.760 (0.752-0.767) | 0.0123 | <0.001 |
| **HRS** | 0.771 (0.761-0.781) | 0.753 (0.742-0.764) | 0.0178 | <0.001 |
| **ELSA** | 0.800 (0.786-0.814) | 0.793 (0.779-0.808) | 0.0065 | 0.008 |
| **MHAS** | 0.732 (0.702-0.763) | 0.730 (0.700-0.761) | 0.0018 | 0.389 |
| **CRELES** | 0.661 (0.612-0.710) | 0.629 (0.577-0.682) | 0.0316 | 0.001 |
| **CHARLS** | 0.763 (0.749-0.776) | 0.757 (0.743-0.772) | 0.0052 | 0.003 |
| **By race/ethnicity** |  |  |  |  |
| **White** | 0.780 (0.771-0.789) | 0.764 (0.755-0.774) | 0.0154 | <0.001 |
| **Black** | 0.683 (0.657-0.710) | 0.679 (0.655-0.704) | 0.0041 | 0.307 |
| **Hispanic/Latino** | 0.746 (0.724-0.768) | 0.740 (0.718-0.762) | 0.0059 | 0.09 |
| **Asian** | 0.763 (0.750-0.776) | 0.756 (0.741-0.770) | 0.0071 | <0.001 |
| **By sex** |  |  |  |  |
| **Men** | 0.757 (0.746-0.767) | 0.745 (0.734-0.757) | 0.0113 | <0.001 |
| **Women** | 0.788 (0.778-0.797) | 0.774 (0.764-0.785) | 0.0133 | <0.001 |
| **By number of comorbidities** |  |  |  |  |
| **0** | 0.779 (0.761-0.796) | 0.767 (0.748-0.786) | 0.0117 | 0.001 |
| **1** | 0.758 (0.744-0.773) | 0.747 (0.730-0.763) | 0.0115 | <0.001 |
| **≥2** | 0.722 (0.711-0.732) | 0.713 (0.702-0.724) | 0.0083 | <0.001 |

**Supplementary Table 3.**
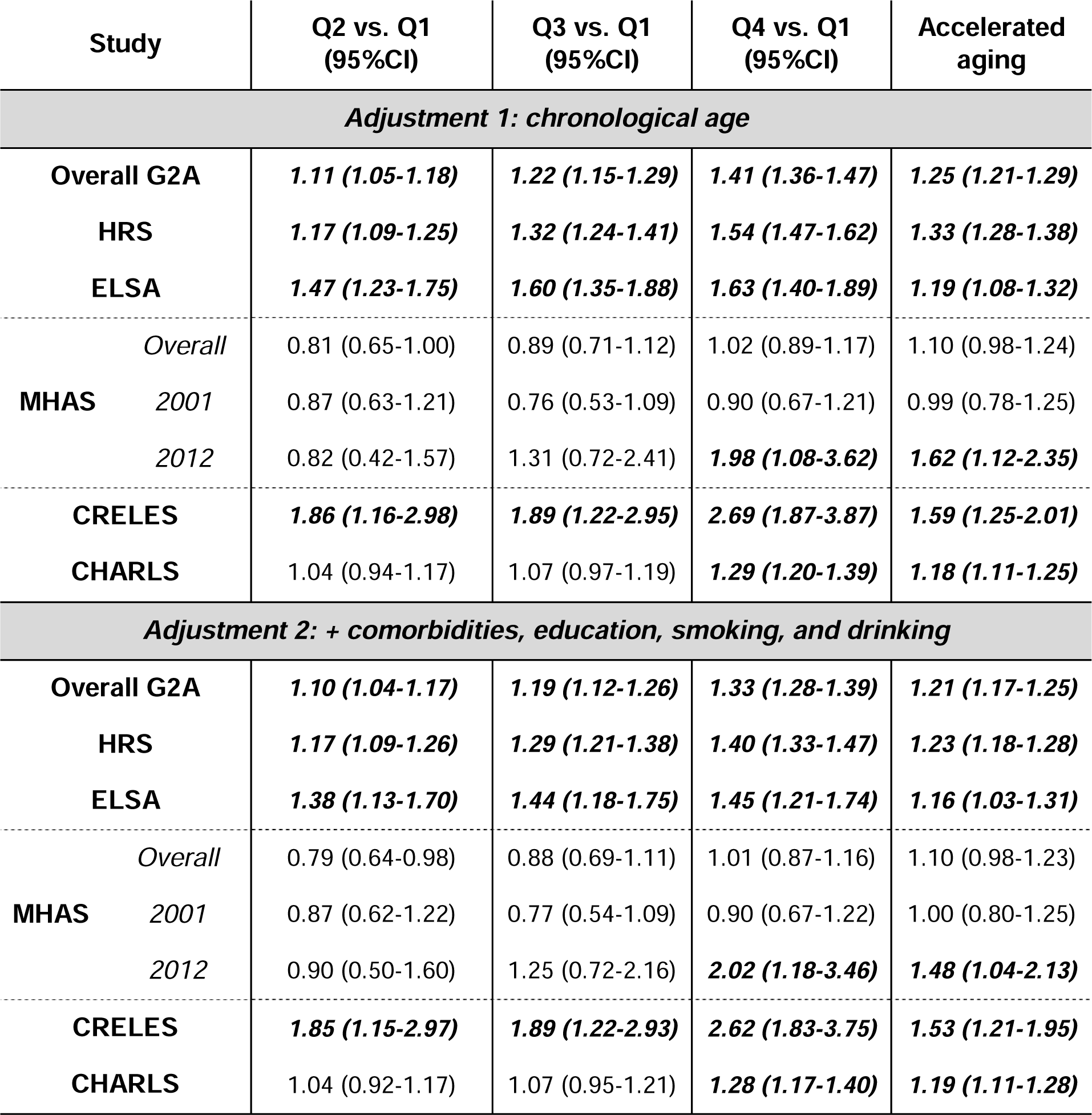
Hazard ratios (95% CI) extracted from Cox proportional hazard regression models clustered by ID (which accounts for repeated measures over time). All-cause mortality risk was compared across AnthropoAgeAccel quartiles and across participants with and without accelerated aging (AnthropoAgeAccel values ≥0). Models were stratified by sex and race/ethnicity and progressively adjusted for 1) chronological age, and 2) number of comorbidities, education and lifestyle.

## SUPPLEMENTARY FIGURES

**Supplementary Figure 1.**
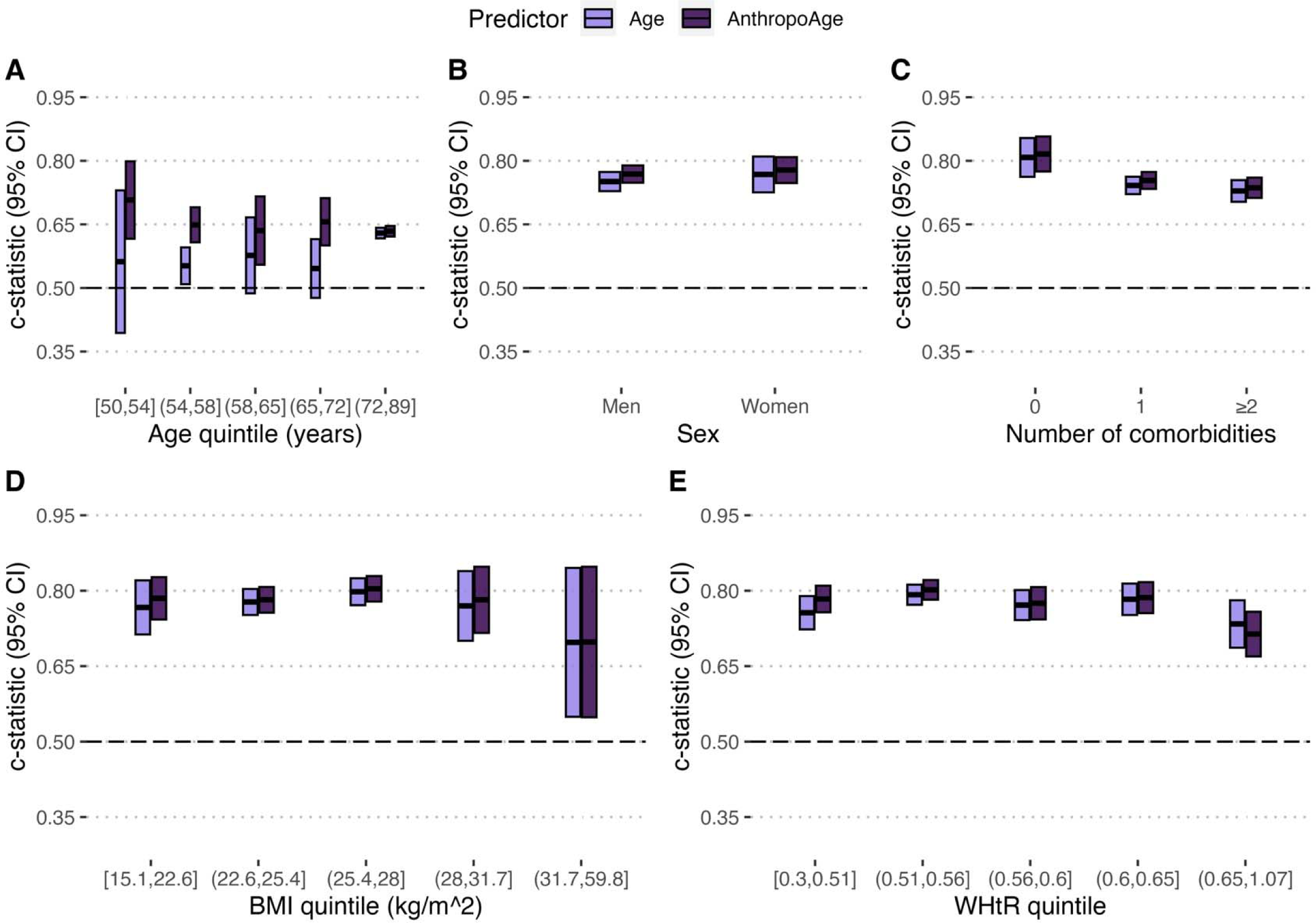
Uno’s c-statistic for prediction of all-cause mortality in the overall G2A population using Cox regression stratified by sex and ethnicity. Results are presented according to chronological age quintiles (**A**), sex (**B**), number of comorbidities (**C**), body-mass index (BMI) quintiles (**D**) and waist-to-height ratio (WHtR) quintiles (**E**).

**Supplementary Figure 2.**
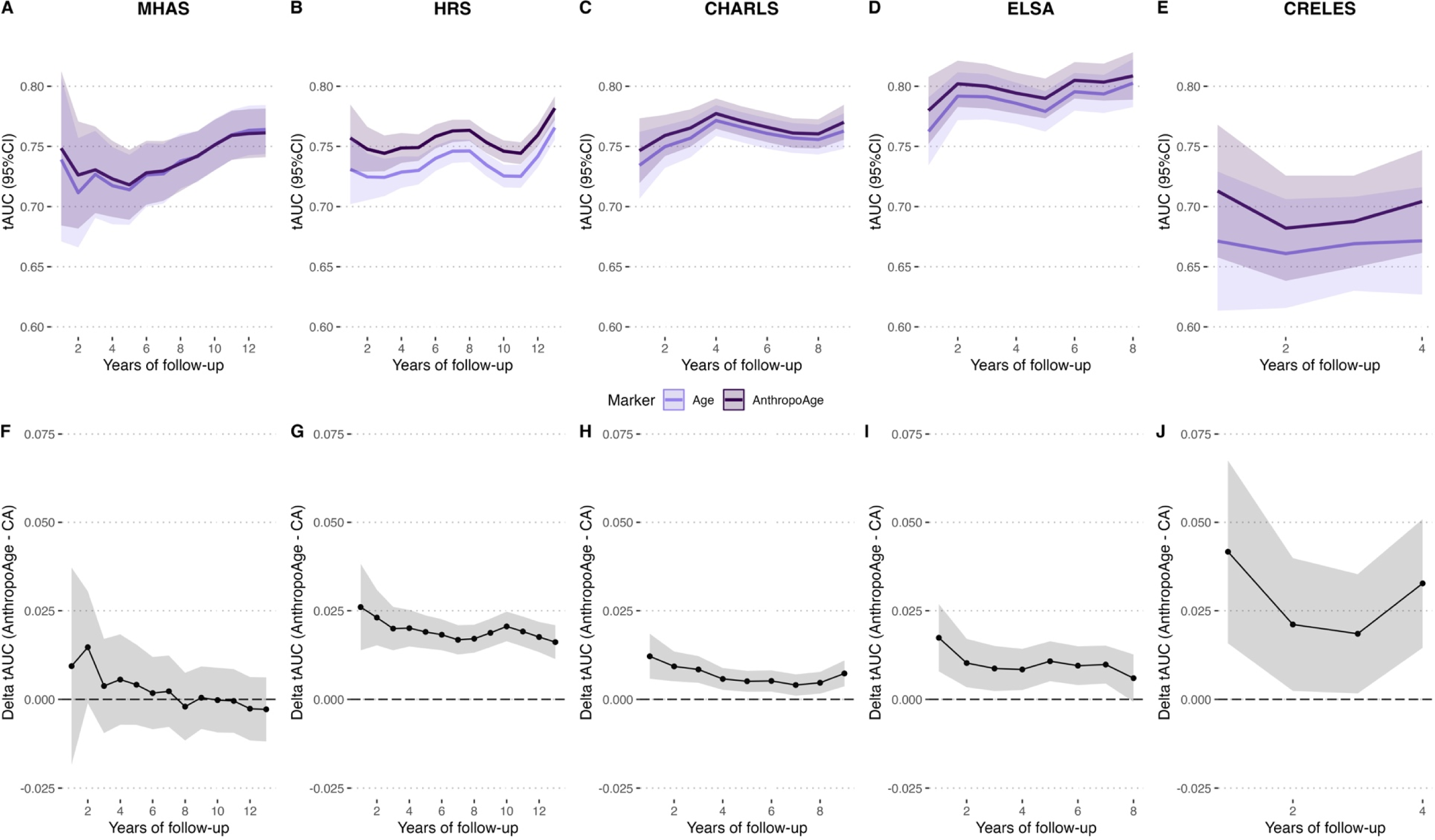
Time-dependent area under the receiving operating characteristic curves (tAUC) evaluated using inverse probability of censoring weights for prediction of all-cause mortality comparing AnthropoAge versus chronological age (CA) in MHAS (assessed over 12 years) (**A**), HRS (12 years) (**B**), CHARLS (8 years) (**C**), ELSA (8 years) (**D**), and CRELES (4 years) (**E**), studies are arranged by length of follow-up. The differences in tAUC (delta tAUC) between AnthropoAge and CA is displayed below (**F-J**).

**Supplementary Figure 3.**
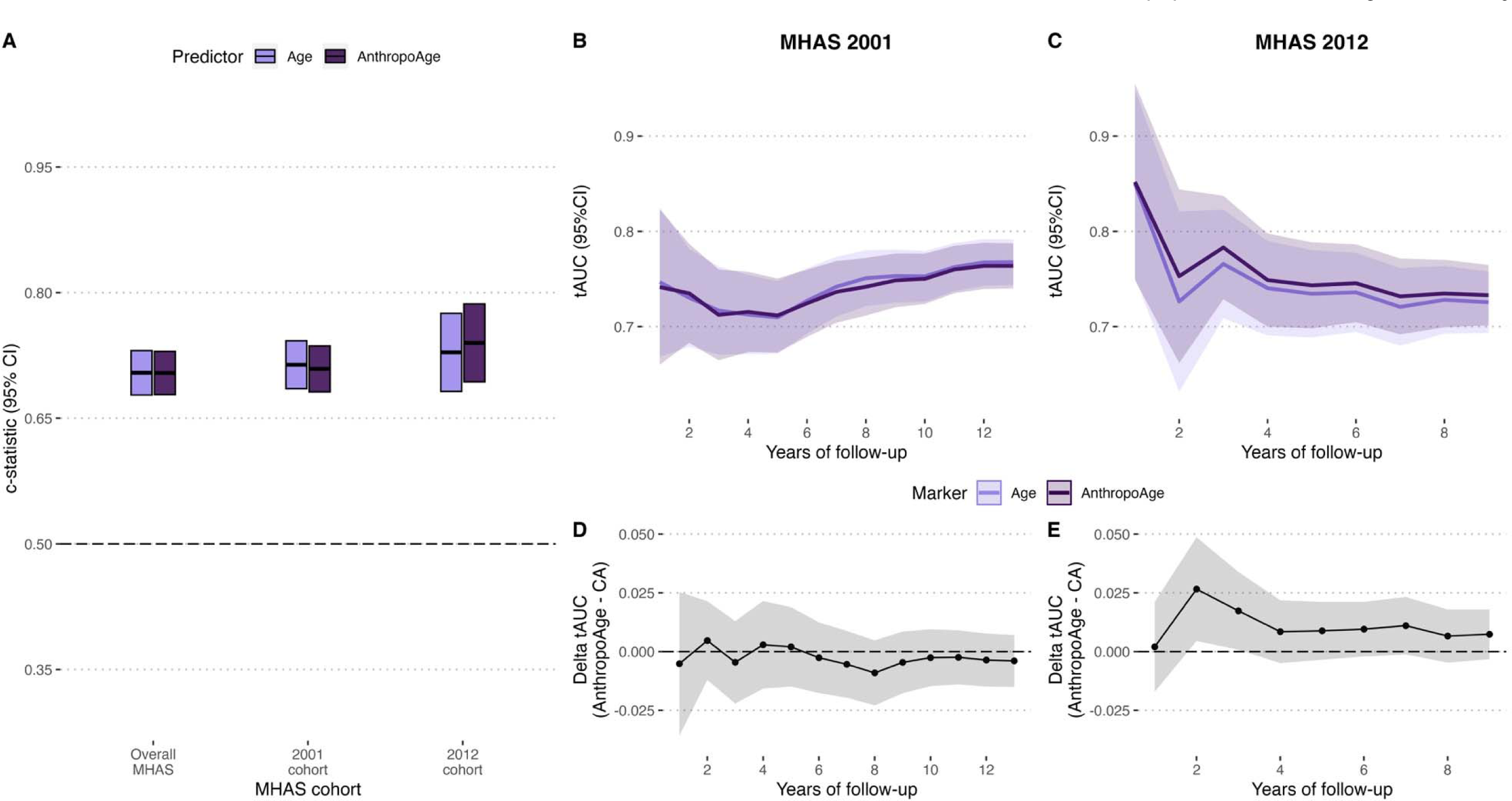
The overall MHAS study comprises a first cohort than began follow-up in 2001 and a second cohort that began follow-up in 2012 with significant differences regarding representativity of anthropometric measurements. Here, we show Uno’s c-statistic comparing the overall MHAS, MHAS 2001, and MHAS 2012 cohorts (**A**); as well as time-dependent area under the receiving operating characteristic curves (tAUC) for MHAS 2001 over 12 years of follow-up (**B**) and MHAS 2012 over 8 years of follow-up (**C**), along with their respective delta tAUC comparing AnthropoAge versus CA (**D-E**).

**Supplementary figure 4.**
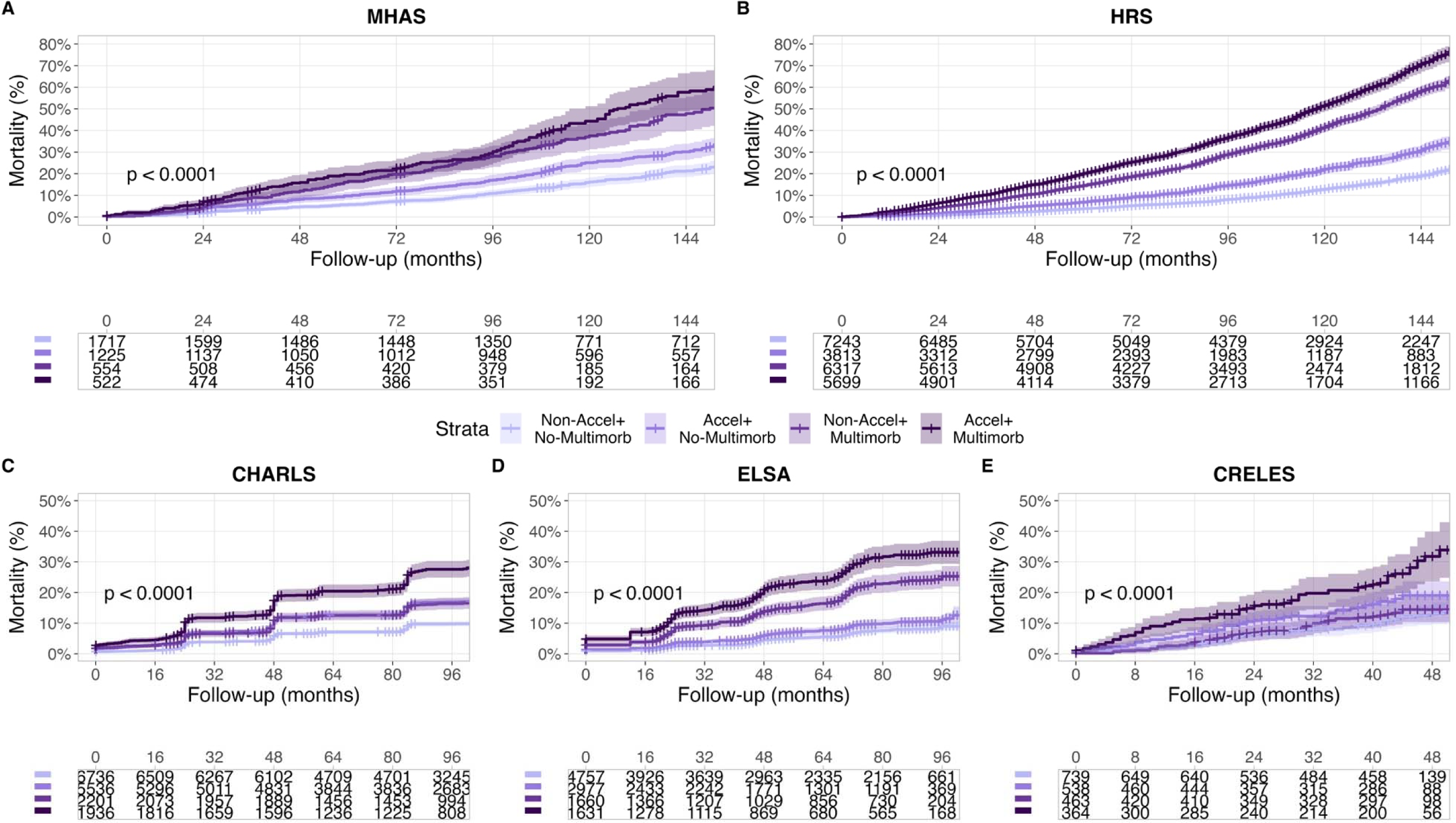
Kaplan-Meier curves comparing cumulative mortality risk of individuals with and without accelerated aging (AnthropoAgeAccel ≥0) and with or without of multimorbidity (≥2 comorbidities) for each study separately. Studies were arranged by follow-up time: 12 years for MHAS (**A**) and HRS (**B**), 8 years for CHARLS (**C**) and ELSA (**D**), and 4 years for CRELES (**E**). Shown p-values are from log-rank tests for comparison of survival curves.

**Supplementary Figure 5.**
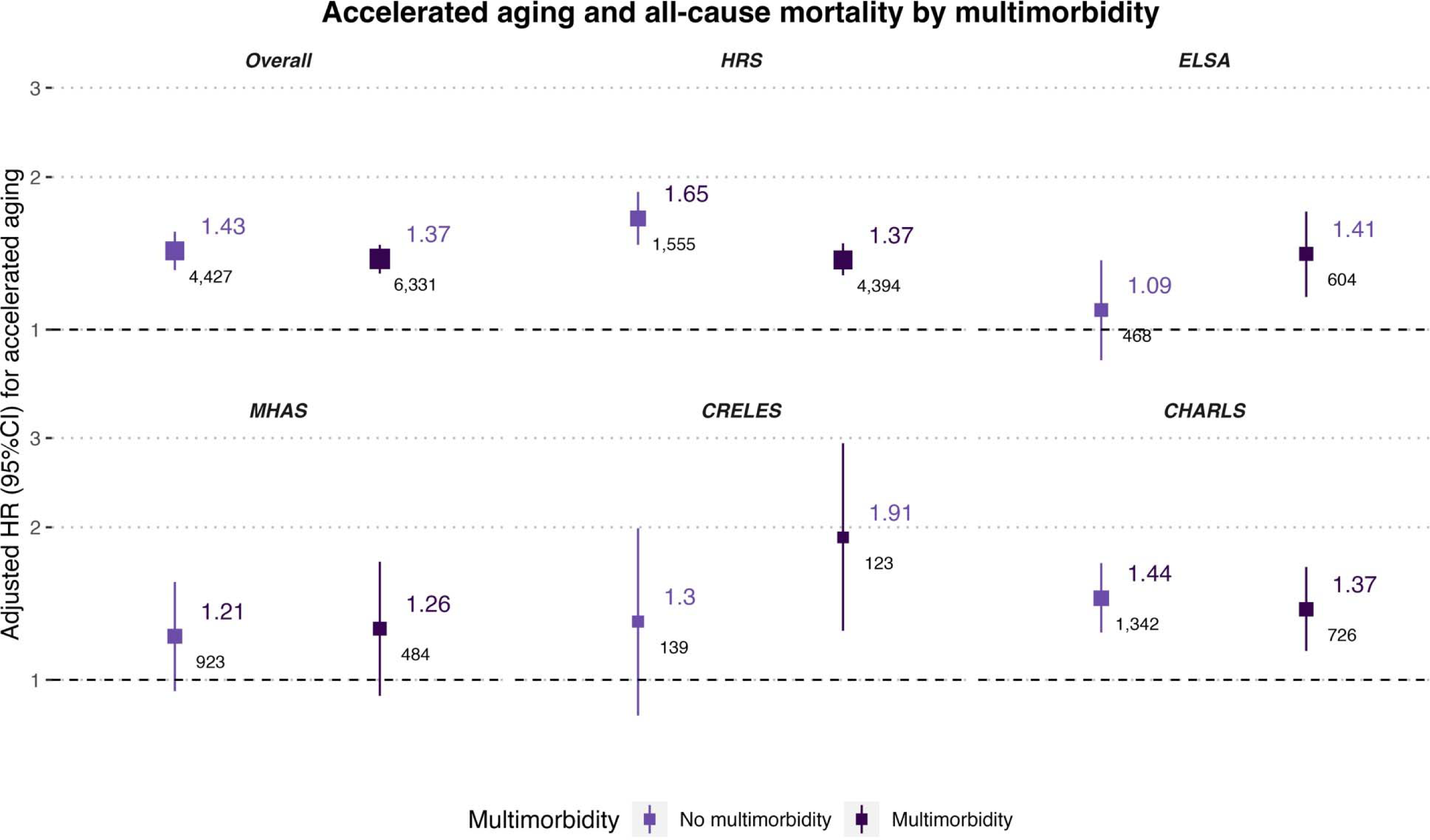
Adjusted hazard ratio (95% CI) plots of all-cause mortality predicted by accelerated aging, obtained from Cox proportional hazard regression models adjusted by chronological age, and stratified by sex, and race/ethnicity. Estimates were obtained separately for participants with and without multimorbidity (≥2 comorbidities). The larger top numbers represent adjusted hazard ratios, and the smaller numbers represent the number of deaths in each group.

**Supplementary Figure 6.**
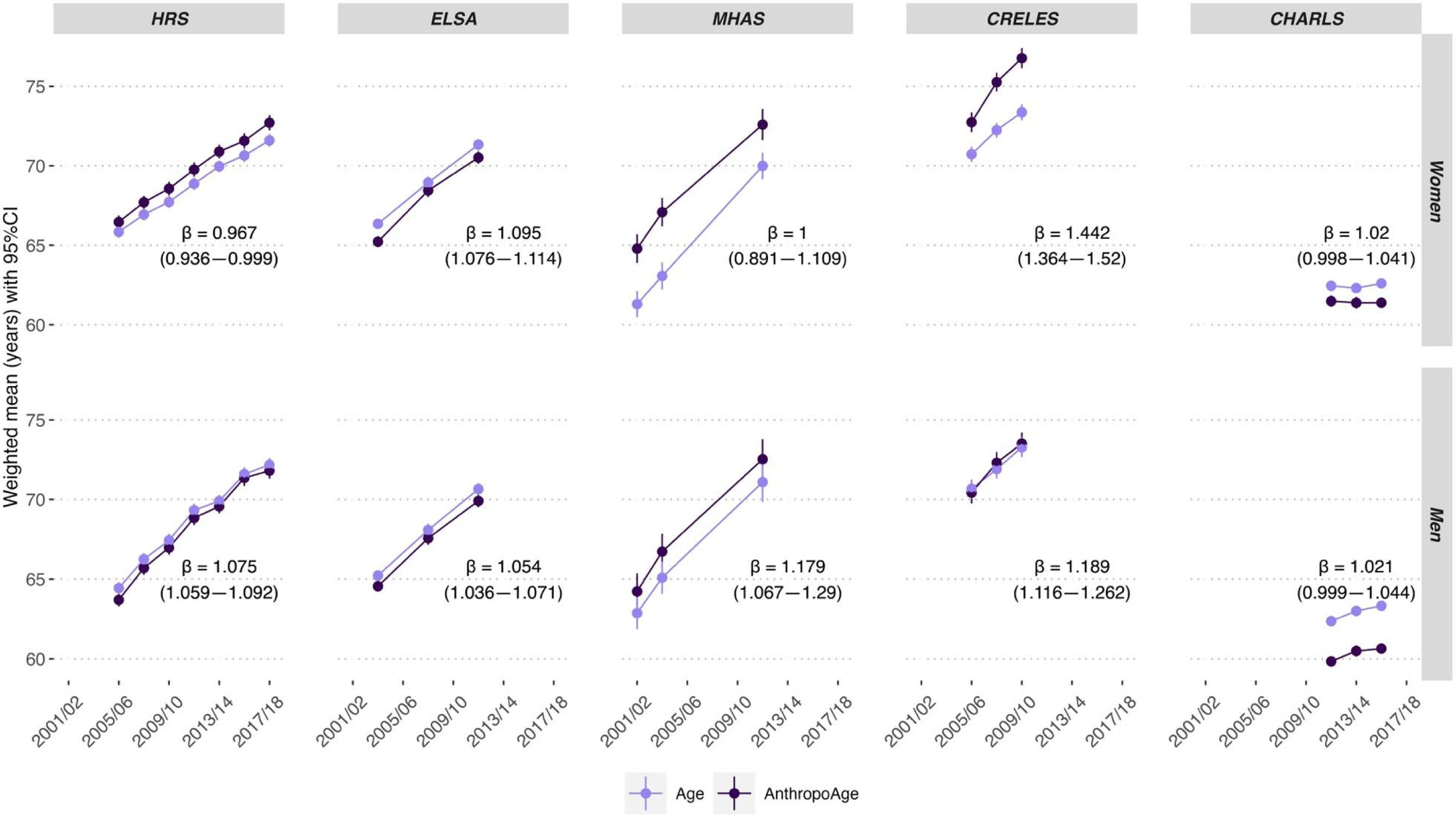
Trends of weighted mean (95% CI) chronological age compared to mean AnthropoAge over time for each G2A study, ***stratified by sex***. A β-coefficient >1 indicates that the rate of population aging occurs, on average, faster than expected with each year of follow-up; β-coefficients were derived from GEE models for individual countries. We only included participants who were present in the first assessment and with a total of at least two visits.

**Supplementary Figure 7.**
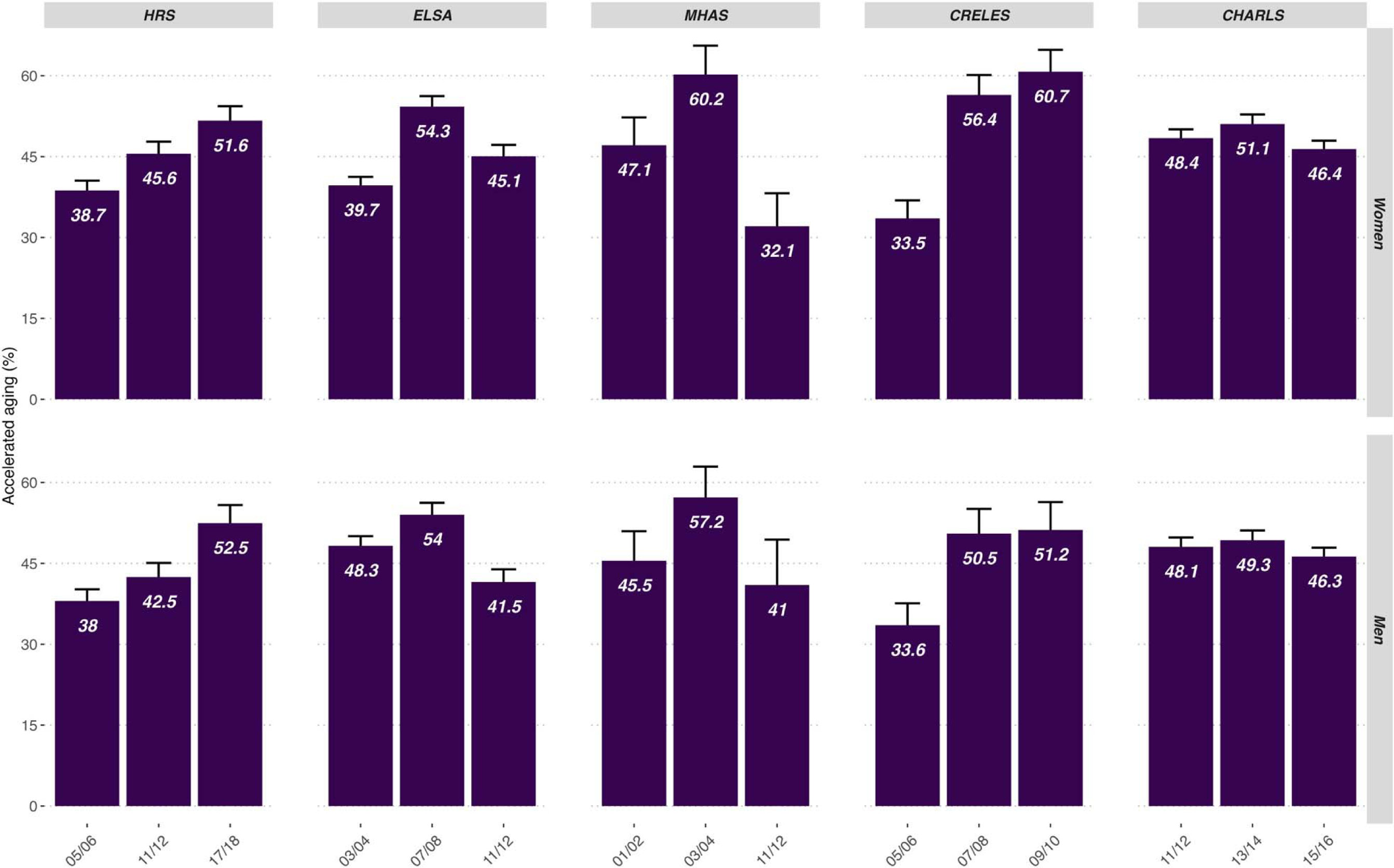
Bar graphs showing weighted prevalence of accelerated aging (with 95% CI) defined as AnthropoAgeAccel ≥0 in biennial cycles for each G2A study, stratified by sex.

**Supplementary Figure 8.**
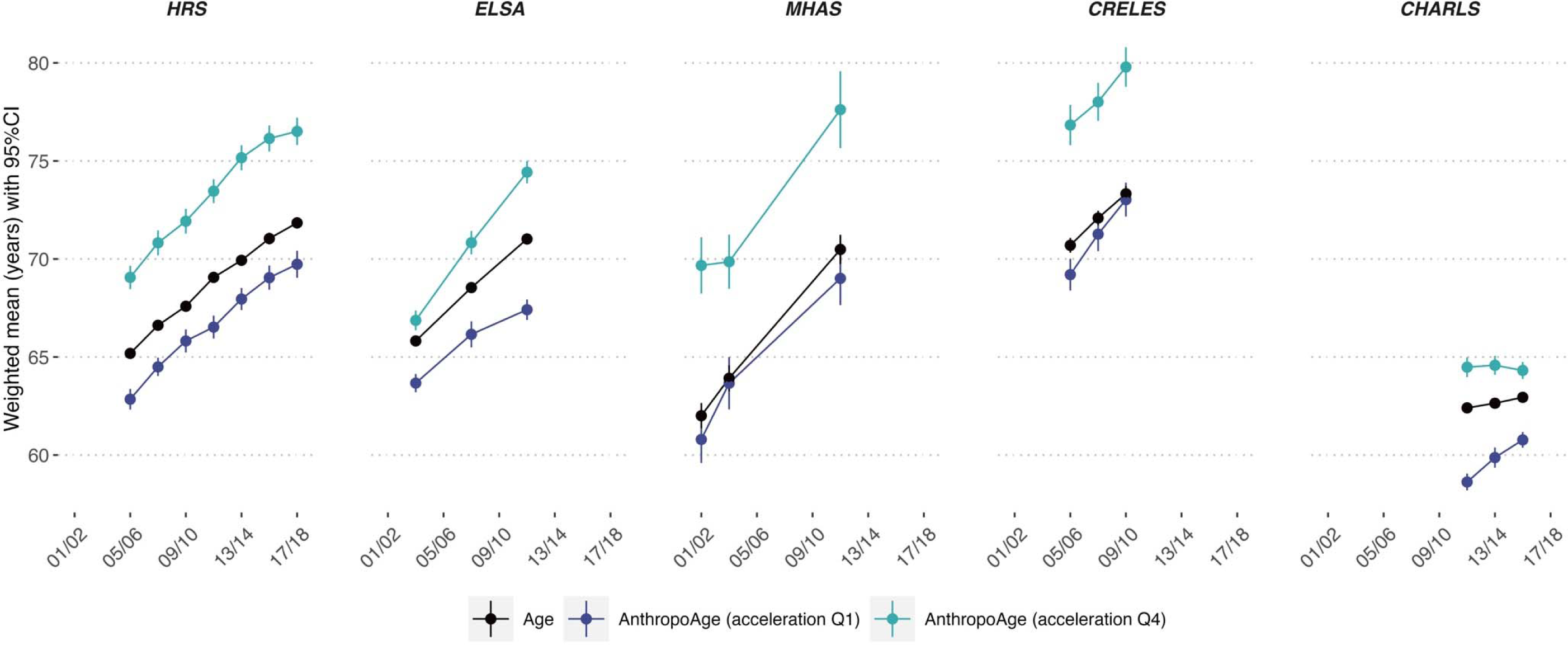
Trends of weighted mean (95% CI) chronological compared to mean AnthropoAge over time for each G2A study. Age values represent the whole population, while AnthropoAge values were stratified for individuals in the ***lower (Q1) vs. upper (Q4) AnthropoAgeAccel quartiles***. A β-coefficient >1 indicates that the rate of population aging occurs, on average, faster than expected with each year of follow-up; β-coefficients were derived from GEE models for individual countries.

**Supplementary Figure 9.**
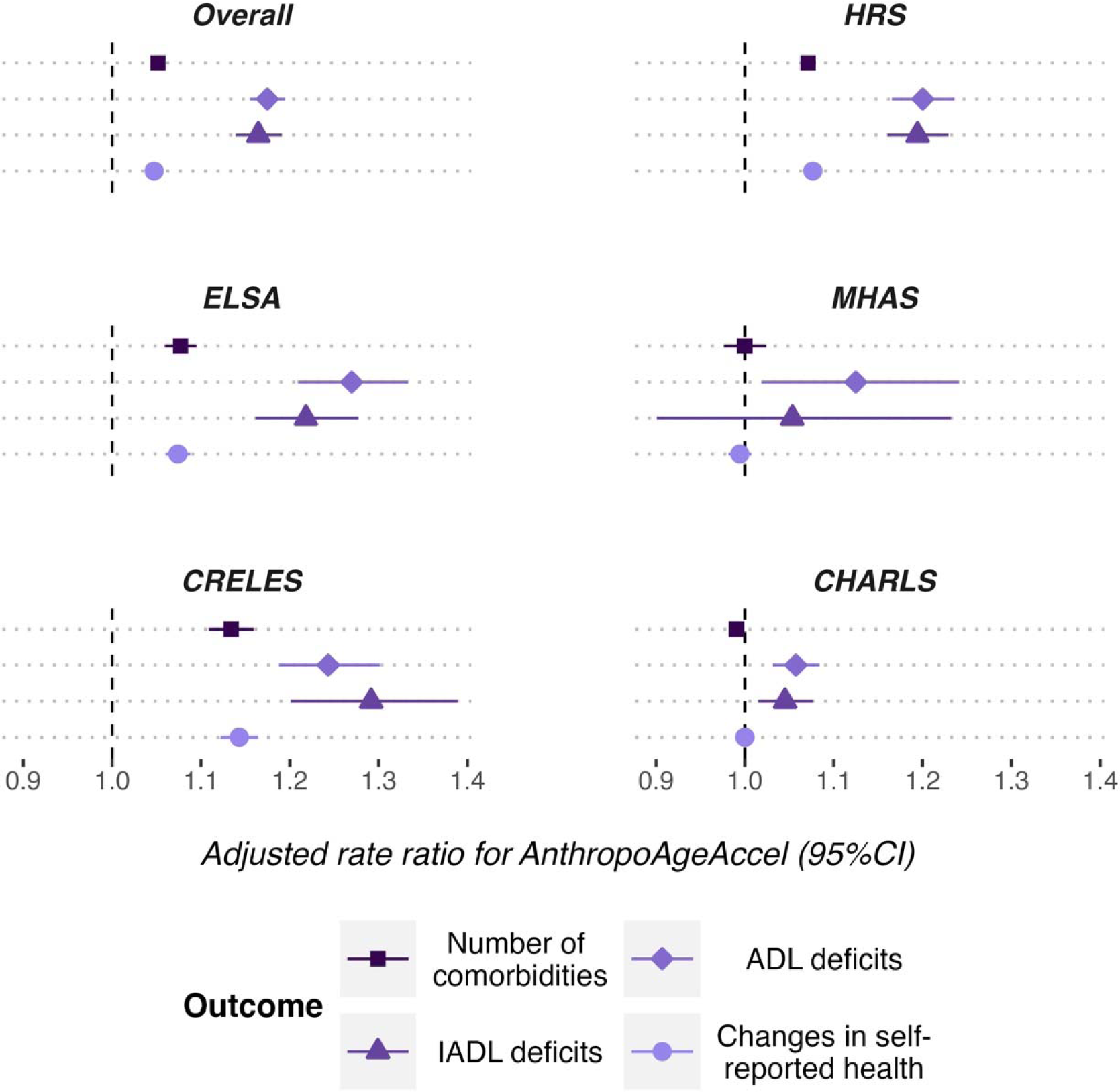
Adjusted rate ratio (95% CI) plots for the association of AnthropoAgeAccel with changes in self-reported health, ADL/IADL deficits and number of comorbidities across G2A studies modeled with GEEs with a Poisson variance function. In this sensitivity analysis, we did not exclude participants who already presented with the outcome at baseline.

